# Impact of human movement between hypo- and hyperendemic areas on sustainability of elimination of *Onchocerca volvulus* transmission

**DOI:** 10.1101/2023.01.20.23284850

**Authors:** Karen McCulloch, Shannon M. Hedtke, James McCaw, Jodie McVernon, María-Gloria Basáñez, Martin Walker, Annette C. Kuesel, Warwick N. Grant

**Affiliations:** Department of Environment and Genetics, La Trobe University, Bundoora, Victoria, Australia; Department of Infectious Diseases, Melbourne Medical School, University of Melbourne at The Peter Doherty Institute for Infection and Immunity, Parkville, Victoria, Australia; School of Mathematics and Statistics, The University of Melbourne, Parkville, Victoria, Australia; Centre for Epidemiology and Statistics, Melbourne School of Population and Global Health, The University of Melbourne, Parkville, Victoria, Australia; Infection Modelling, Murdoch Children’s Research Institute, Melbourne, Victoria, Australia; MRC Centre for Global Infectious Disease Analysis and London Centre for Neglected Tropical Disease Research, Department of Infectious Disease Epidemiology, School of Public Health, Imperial College London, London, United Kingdom; UNICEF/UNDP/World Bank/World Health Organization Special Programme for Research and Training in Tropical Diseases (TDR), World Health Organization, Geneva, Switzerland

## Abstract

**Background:** Onchocerciasis is a vector-borne disease caused by the filarial nematode *Onchocerca volvulus*. Endemic countries target elimination of parasite transmission using primarily annual ivermectin mass administration. Elimination is particularly challenging in sub-Saharan Africa, where there are large contiguous areas with varying levels of endemicity and intervention history. We examined one challenge to elimination that has received little attention to date: movement of people between areas.

**Methodology/Principal Findings:** We extended one of the commonly used onchocerciasis transmission models, EPIONCHO, to allow modelling the effect of movement of people and/or flies between areas (“patches”). We explored the impact of humans travelling between a hypoendemic area (i.e., with low vector biting rates) with no history of interventions and a hyperendemic area (high vector biting rates) that stopped intervention (ivermectin mass administration) after infection prevalence decreased below 1.4%. Our results suggest that human travel in either direction will accelerate recrudescence in the hyperendemic area to pre-intervention levels, relative to recrudescence without travel, and can reduce the annual biting rate required for parasite transmission within hypoendemic areas.

**Conclusions/Significance:** Our results emphasize the importance of decisions on which hypoendemic areas to include in interventions and suggest that transmission mediated by human movement needs to be considered when planning (a) the geographic areas and sampling density for evaluations for decisions on when and where to stop interventions, (b) where, how often, and for how long to conduct post-intervention surveillance for verification of elimination of transmission and (c) where and how frequently to conduct post-elimination surveillance. Given the cost implications of stopping interventions too early or later than necessary, we encourage the development of models such as the one presented here for quantitating the impact of human and vector movement between areas on the risk and timeframe of recrudescence after interventions are stopped to inform economic analyses.

**Author summary:** Onchocerciasis is an infectious parasitic disease that causes significant morbidity, from incessant skin itching to blindness. Onchocerciasis has also been implicated as the cause of high epilepsy rates. Efforts are underway to eliminate the parasite. Mathematical models for parasite transmission between humans via blackflies can be used to explore how interventions (e.g., mass drug administration or blackfly control) impact the percentage of people infected (infection prevalence). We extended a commonly used model to explore how people travelling between areas that differ in blackfly abundance, infection prevalence, and past interventions affects infection prevalence. We found that people travelling between an area with few blackflies (hypoendemic), low infection prevalence, and no interventions and an area with many blackflies (hyperendemic) but very low infection prevalence thanks to many years of mass drug administration which was then stopped will accelerate the increase in infection prevalence in the hyperendemic area compared to a situation where no people travel between these areas. This means that strategies for onchocerciasis elimination need to consider the effect of humans travelling.

## Introduction

Onchocerciasis is a neglected tropical disease caused by the filarial nematode *Onchocerca volvulus* transmitted by *Simulium* (blackfly) vectors [1-3]. The prevalence of infection and morbidity and the resulting socio-economic impact have motivated large-scale control and elimination programs in affected countries in Central and South America, Sudan, Yemen, and sub-Saharan Africa. In 2020, around 240 million people were estimated to require inclusion in these programs, with >99% in sub-Saharan Africa [4]. The principal intervention is currently annual, and in some cases biannual, mass drug administration of ivermectin (MDAi).

For many years, it was assumed that MDAi could eliminate *O. volvulus* transmission in the small endemic areas in the Americas and possibly Yemen, but not across sub-Saharan Africa [5]. However, research and epidemiological evaluations indicated that MDAi reduced prevalence of infection in many areas in Africa more effectively than anticipated [6-9]. This has resulted in African endemic countries now targeting not only control of onchocerciasis as a public health problem, but elimination of parasite transmission [10, 11]. This shift requires re-evaluation of current activities and tools used for determining where, and for how long, which interventions need to be deployed, and subsequent development of new tools, activities, and strategies where needed [3, 12-23].

The impact of the highly heterogeneous prevalence of onchocerciasis in sub-Saharan Africa [24-26] on persistence of transmission needs to be incorporated into this decision making. Areas pre-MDAi were categorized based on estimated subcutaneous nodule prevalence as hypoendemic, defined in countries working with the African Programme for Onchocerciasis Control (APOC; 1995-2015) as areas where estimated subcutaneous nodule prevalence was <20% (corresponding, with wide variation, to a skin microfilariae [mf] prevalence of <35% [27-30], mesoendemic (nodule prevalence 20-40%; approximate mf prevalence 35-60%), and hyperendemic (nodule prevalence >40%; mf prevalence >60%). Importantly, while onchocerciasis control as a public health problem was targeted, onchocerciasis hypoendemic areas were included in MDAi only when they belonged to a health system administrative unit which included meso- and hyperendemic areas [31, 32]. While the source of infections in hypoendemic areas may be nearby meso- or hyperendemic areas, modeling has demonstrated plausible mechanisms for stable and self-sustaining hypoendemic transmission [33]. To achieve sustainable elimination of transmission, we need to understand the risks hypoendemic areas pose to elimination in the absence of interventions. and develop tools for national programs to determine where interventions are needed and where and when to stop MDAi (and/or other interventions, if applicable). Furthermore, we need to delineate the geographic areas which belong to an ‘endemic onchocerciasis focus’ that need to be included in evaluations for stop MDAi decisions and to determine where, how frequently, and for what duration to carry out post-MDA surveillance as well as surveillance after elimination of transmission has been verified by WHO.

Mathematical modeling of *O. volvulus* transmission and the effect of vector control or MDAi has for many years informed program strategies and objectives [31, 34-41]. This has included estimating the time periods during which countries may meet the APOC-defined *<u>p</u>*rovisional *<u>o</u>*perational *<u>t</u>*hreshold for *<u>t</u>*reatment *i*nterruption and commencement of *<u>s</u>*urveillance (pOTTIS) based on pre-MDAi endemicity, reported MDAi duration, and treatment coverage [31, 42] and identification of barriers for reducing infection prevalence, particularly in meso- and hyperendemic areas (including low treatment coverage (i.e., low proportion of a community taking ivermectin in each round of MDAi) and/or systematic non-compliance (i.e., proportion of individuals never or very rarely taking ivermectin)) [37, 39, 43]. Recently, the ONCHOSIM [34, 36, 38, 39] and EPIONCHO [39, 40, 44-46] models have been extended to incorporate different types of heterogeneity, such as in human host mixing patterns [33] or exposure to blackflies [47], to understand the impact these factors have on elimination efforts. Equally critical to inform country programs is quantifying uncertainty in modeled thresholds [48, 49] or duration of MDAi to achieve elimination [37] due to assumptions about parameter estimates, such as density-dependent regulation of parasite establishment within the human and vector hosts [47, 50]. Modelling is also now addressing challenges for decisions on where and when to stop MDAi so that elimination is sustained, including the appropriateness of the criteria and procedures for stopping MDAi in the WHO 2016 guidance and the impact of strategies other than MDAi [3, 12, 50-56].

The impact of heterogeneity over different geographic scales in endemicity and intervention history and the role of human and vector movement on transmission on elimination efforts remain largely unexplored. This includes the potential for re-introduction of parasites through human or vector movement from areas with ongoing transmission into areas where interventions were stopped after transmission was considered to have been eliminated [57-59]. Long-range seasonal vector migration was already recognized as an important challenge for interrupting transmission through vector elimination by the Onchocerciasis Control Programme in West Africa (OCP, 1974-2002) [60-65]. People travel within and across countries for many reasons, including, but not limited, to work, visiting family, or attending ceremonies and celebrations. This may impact parasite transmission. In this context, the exclusion of hypoendemic areas from MDAi while control of onchocerciasis as a public health problem, not elimination of transmission was targeted, has prompted several questions, including (i) what sustains low levels of infection in these areas, i.e., is transmission ongoing within these areas or is there ongoing import of infections from meso- or hyperendemic areas? and (ii) do these areas pose a risk for recrudescence in areas which have met MDAi cessation criteria and discontinued MDAi?

Modeling studies have been utilized to address question (i), and illustrated that sustained transmission in isolated hypoendemic areas could be attributed to increased assortative mixing within the human host population, i.e., when individuals with similar exposure to blackfly bites are more connected to each other [33]. They showed that onchocerciasis transmission could not be sustained below 35% mf prevalence if the human population was assumed to be homogenously mixing given moderate levels of heterogeneity in exposure to vectors. However, when the human population was assumed to have heterogeneous exposure to blackflies with assortative mixing, onchocerciasis transmission could be sustained at mf prevalence levels as low as 8%. The recent extension of the ONCHOSIM model to consider spatial heterogeneity in transmission of *O. volvulus* between areas of different endemicity and intervention history demonstrated that in hypoendemic settings with an annual biting rate (ABR) below the threshold required to sustain transmission of *O. volvulus*, transmission can persist if there is regular movement of infected humans or infective blackflies from areas of higher ABR [56]. The authors concluded that expanding MDAi into hypoendemic areas would shorten the duration of MDAi required to achieve elimination in nearby meso- and hyperendemic areas ‘connected’ to the hypoendemic areas through human movement.

In this paper, we focus on addressing question (ii) to understand the risk hypoendemic areas pose for recrudescence in nearby hyperendemic areas which have discontinued MDAi. We introduce a deterministic patch model framework describing the transmission of *O. volvulus* in areas of different levels of endemicity and/or MDAi history and assessing the impact that human and blackfly movement between these areas has on the prevalence of infection. We utilize this model to explore the impact of humans moving between a hypoendemic area, where MDAi was never implemented, and a hyperendemic area, where MDAi was discontinued (illustrated in Fig 1) on infection prevalence and the annual transmission potential (ATP) in both areas and thus the risk and time-frame of recrudescence in the post-MDAi hyperendemic area and sustainability of local transmission in the hypoendemic area.

**Figure 1.**
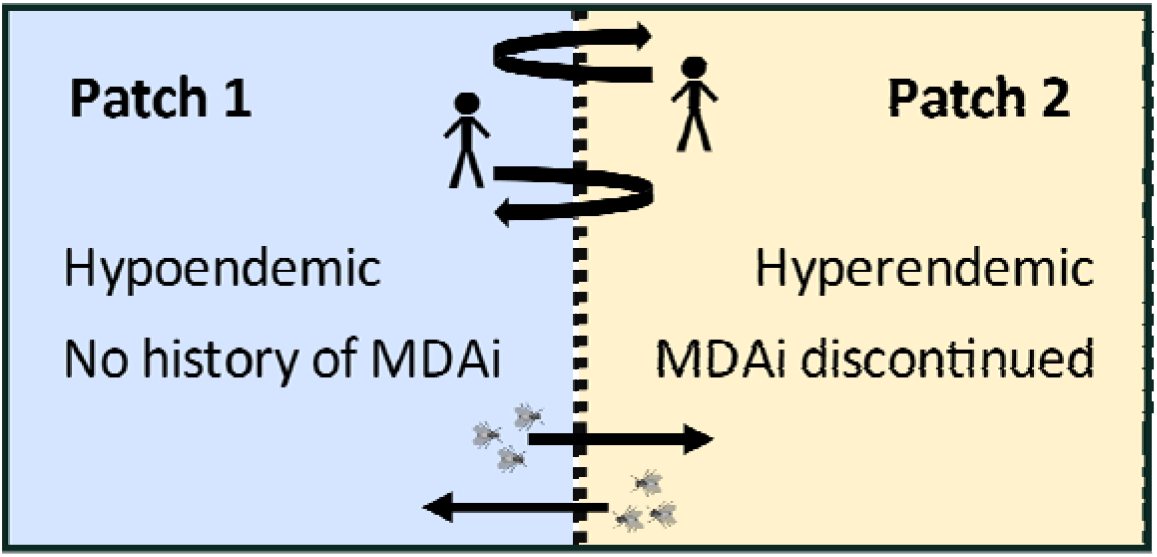
Illustration of modeled movement between two areas (patches) with different endemicity and control history. Human movement is modeled as commuting and vector movement as migration.

## Methods

The population-based deterministic model EPIONCHO [37, 39, 40, 44, 46] describes the transmission of *O. volvulus* within a single homogeneously mixing population. EPIONCHO is an “intensity” compartmental model. Unlike epidemic compartmental models (e.g. [66]) where the number of susceptible and infected hosts are each tracked, intensity models track the average parasite intensity within the human and vector host populations. Intensity models that track the distribution of parasites within the human and vector host populations are commonly used to model macroparasitic diseases, including lymphatic filariasis [67-69] and onchocerciasis [34, 38-40, 44, 46].

Patch models, sometimes referred to as metapopulation models, have been utilized to model the spread of infection where transmission between populations or areas is important. In a patch model, the population of interest is divided into distinct subpopulations (or patches), where each patch has distinct characteristics such as population size and disease transmission dynamics [66, 70]. An illustration of a 2-patch model is shown in Figure 1 and Figure 2.

**Figure 2.**
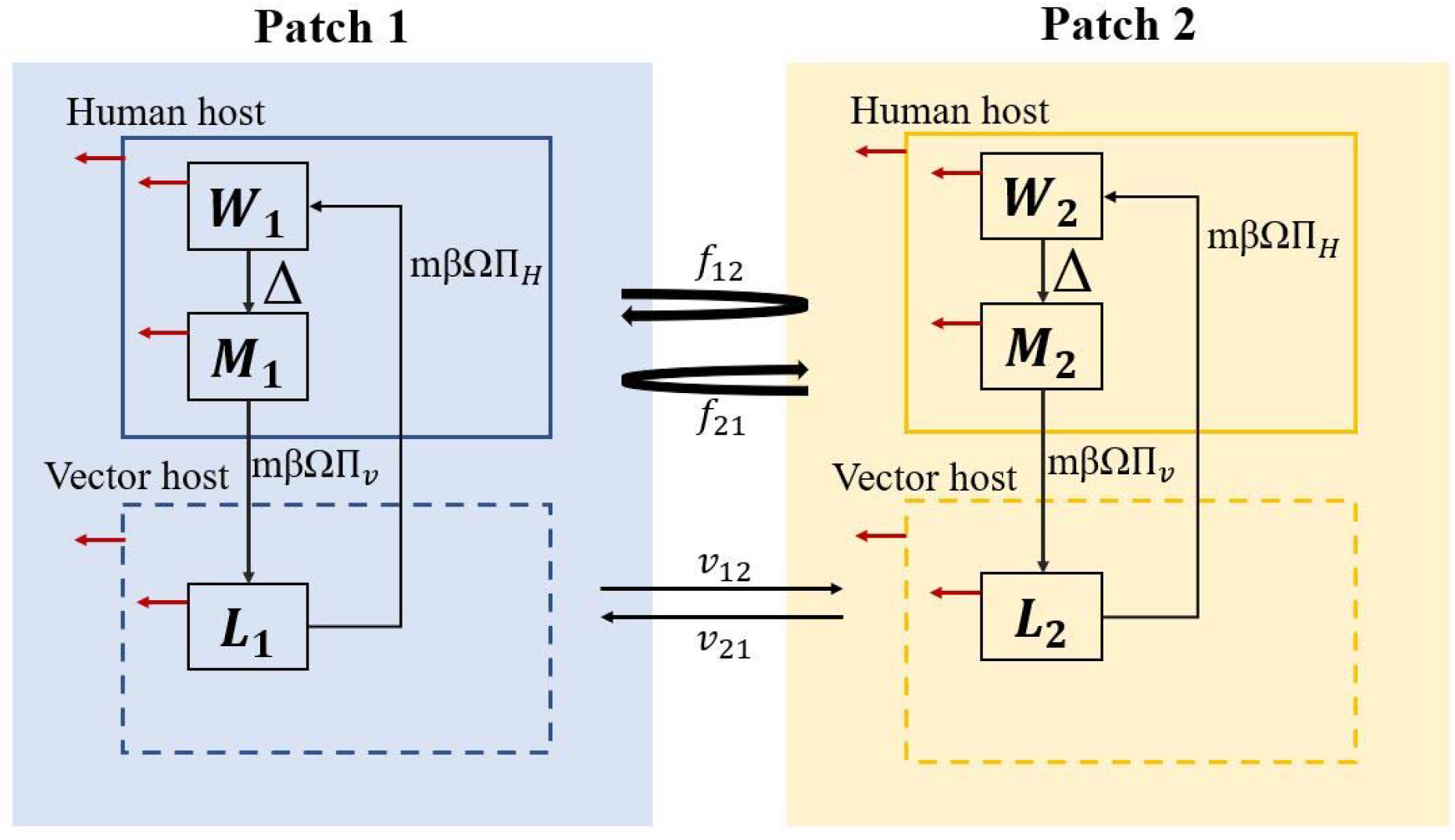
Schematic diagram of a 2-patch intensity model. *W*_*i*_ is the mean number of adult worms per human host in patch *i, M*_*i*_ is the mean number of microfilariae per mg of skin within the human host in patch *i*, and *L*_*i*_ is the mean number of infective larvae per blackfly in patch *i*. Red arrows represent mortality.

EPIONCHO as described by Filipe et al. [46] is an age- and sex-structured model for the intensity of *O. volvulus* infection that incorporates parasite regulation within the humans and vector hosts, in addition to heterogeneity in exposure to blackfly bites. This model provides a realistic model framework for the transmission dynamics of *O. volvulus* (later expanded to incorporate interventions such as MDAi or vector control [71]). Here, we extended this framework into a patch model to describe the impact of human and vector movement on the transmission of *O. volvulus* between numerous areas (patches) of different endemicity (determined by the density of vectors) and MDAi history, resulting in different prevalence and intensity of infection in humans and vectors.

### Model equations and parameters

The intensity of *O. volvulus* infection changes over time and with host age, as described by the system of partial differential equations (PDEs) S.1 – S.8 in Table S1 in S1 File for a 2-patch model. The model describes, with respect to time () and host age (), the number of adult worms in human host of sex resident in patch (), the number of mf in the skin of human hosts of sex resident in patch (), and the number of infective larvae in the blackfly population in patch biting human hosts of sex (). Table S2 in S1 File provides a detailed description of the variables and parameters in the patch model and code. Parameter estimates utilized in this study are appropriate for pre-control savannah settings based on previous studies (most of which were conducted in Cameroon) [40, 46]. The ranges of parameter estimates considered in the sensitivity analysis are provided in Table S2 in S1 File and were determined by the parameter estimates relevant for savannah settings available in published literature. Table S3 in S1 File provides the PDEs for a model with any number of patches ≥2.

### Modeling of human and vector movement

Fig. 2 illustrates how human and vector movement are modeled. To model movement of humans between the patches in both directions (referred to in patch model literature as ‘commuting’), we define as the fraction of time residents of patch in age group spend visiting the connected patch *j* included in the model. The fraction of the population resident in patch *i* of age group *a* that travel is given by and the proportion of time residents of patch that travel spend in patch is given by. For the 2-patch model presented here, *d*_*ij*_ = 1 as there is only one patch for residents to visit. We hypothesize that mobility for work is the dominant reason for travel between the patches, and thus we make the simplifying assumption that only human hosts aged between 20 and 50 years travel, representing 35% of the total population. Therefore, the fraction of time human hosts resident in patch *i* aged 20 to 50 years spend visiting patch *j* is given by *f*_*ij*_. The model defines vector movement between patches as migration, which means once the vectors leave their ‘resident’ patch they do not return. Therefore, we define the parameter *v*_*ij*_ as the migration rate per year of blackflies from patch *i* to patch *j*. Furthermore, we assume that the ratio of the size of the vector population to human population (*m*_*i*_) in patch *i* is constant. The parameter *m*_*i*_ depends on the ABR and the biting rate per fly on humans (*β*) (see Table S1 in S1 File), and therefore will differ between patches with different ABR.

### Endemicity of the patches included in the model

Areas where *O. volvulus* is transmitted are, in most cases, categorized as hypo-, meso-, and hyperendemic based upon the approximate prevalence of infection within the human population prior to implementation of control programs. The prevalence ranges defining these categories differ slightly between references, driven by (for example) increasing knowledge of correlates of morbidity or the method used to estimate infection prevalence (e.g. prevalence of individuals with microscopically detected mf in skin snips, prevalence of individuals with subcutaneous onchocercal nodules) [24, 30, 32, 72, 73].

While the endemicity of areas is usually quantitated via measures of the prevalence of infection in the human population before implementation of interventions, from a biological and modeling perspective, endemicity is determined by the vector prevalence, which is usually quantified by the ABR. From this perspective, an area defined as hyperendemic retains that categorization even if MDAi decreases the skin mf prevalence in a hyperendemic area to <35%, unless vector control (or natural changes) bring about a reduction in ABR. In this context, patch 1 is parameterized to be representative of a hypoendemic area with an ABR of 1000, while patch 2 is parameterized to be representative of a hyperendemic area with an ABR of 4000.

### Scenarios modeled

We used the patch model developed to simulate the effect of human movement between two different areas characterized as follows:

Patch 1 (hypoendemic area). In the absence of past or present control measures, transmission is stable and results in *28%* of the human population over all ages infected. The initial conditions for patch 1 were determined by first simulating an isolated patch 1 (i.e., no movement between patch 1 and 2) to equilibrium.

Patch 2 (hyperendemic area). We assumed that patch 2 had undergone many years of MDAi which was discontinued when the epidemiology-based provisional threshold (pOTTIS) had been reached.

The provisional thresholds were defined by APOC in its “Conceptual and Operational Framework of Onchocerciasis Elimination with Ivermectin Treatment,” which also specifies the plan to review and refine thresholds as more evidence becomes available [74]. The framework was developed in response to the request from APOC’s governing board, the Joint Action Forum, ‘to determine when and where ivermectin treatment can be safely stopped and to provide guidance to countries on preparing to stop ivermectin treatment where feasible’ [13, 31]. In the Conceptual Framework, prevalence estimates are based on the percentage of individuals with skin mf levels detectable via microscopic evaluation. Informed by the experience with stopping interventions in West Africa (vector control in the countries of the OCP and data for the savanna vector species *S. damnosum s*.*s*. and *S. sirbanum* [75] and MDAi in a study in Senegal and Mali [6, 8]), pOTTIS includes an entomological parameter (vector infectivity <0.5 infective flies/1000 flies) and an epidemiological parameters (mf prevalence < 5% in all surveyed villages <u>and</u> < 1% in 90% of those villages 11-12 months after the last MDAi round) [13, 31]. The age range over which the epidemiological parameter is to be applied, the number of skin snips to be taken and duration of skin snip incubation to determine mf prevalence (which impact the sensitivity, [76-78]) are not specified. The surveys conducted in APOC countries after development of the Conceptual Framework to assess progress towards elimination included only children >5 years of age, 2 snips and 24 hours of incubation in saline [9]. In the absence of established transmission breakpoints (infection prevalence or intensity below which transmission is unsustainable and the parasite population will die out [39, 71, 79-81], previous modeling studies parameterized the two-level epidemiological pOTTIS as the weighted mean resulting in <1.4% modelled mf prevalence across *all ages* considered as pOTTIS having been achieved [12, 39, 40, 82]. The epidemiological, rather than the entomological pOTTIS is used since it is reached later and is thus the more conservative threshold [40].

Thus, to model each scenario, patch 2 was initialized with a skin mf prevalence in the human population (across *all ages*) of < 1.4%. As described in Basañéz et al., the pOTTIS does not represent a transmission breakpoint and, in the EPIONCHO framework utilized here, mf prevalence will rebound from the pOTTIS under conditions of high ABR such as found in hyperendemic areas [40]. Thus, what we explore here is the extent to which coupling between two patches (connecting them through human travel between the patches) *accelerates* the modeled rebound in mf prevalence in hyperendemic areas.

We modeled a number of scenarios that differed in the extent of coupling between the two patches, i.e., the extent of human movement from patch 1 to patch 2 or vice versa.

### Model output reported

Consistent with previous work [39, 40], we report the modeled mf prevalence in the human population of those aged ≥ 5 years (subsequently referred to as ‘mf prevalence’). In addition, we report the Annual Transmission Potential (ATP). The ATP is the ‘estimated number of infective *O. volvulus* larvae received by a man exposed for eleven hours a day throughout the year at a particular site’ [83]. The ATP is the arithmetic sum of the Monthly Transmission Potential (MTP) determined for each month of the year. The MTP is the product of the monthly biting rate (MBR, total number of blackflies collected in a month divided by the number of days of capture in this month multiplied by 30) and the total number of infective *O. volvulus* larvae (L3) identified in the heads of dissected flies divided by the total number of flies dissected in that month [59]. Dissection of flies and microscopic identification of L3 in their heads can be replaced by examination of flies (or bodies and heads of dissected flies) pooled by sampling site and date of collection for the *O. volvulus* specific O-150 DNA sequence using polymerase change reaction (O-150 PCR pool-screening, [51, 84, 85]. For calculation of ATP in EPIONCHO see Table S2 in S1 File.

### Sensitivity Analysis

To assess the sensitivity of the modeled long-term mf prevalence in each patch to variations of input parameters, we utilized the Partial Rank Correlation Coefficient (PRCC) method as outlined in Marino et al. [86]. Selected parameters were varied according to the distributions provided in Table S2 in S1 File, which were informed by published literature (Table S2 in S1 File). We performed Latin hypercube sampling (LHS) for 1,000 samples of each parameter, then used the LHS outputs to derive PRCCs. The results of PRCC methods provide a measure of the strength of the relationship between the input parameters and the selected output. In addition, PRCC provides information on how influential each parameter is on the output when compared to other parameters, where a PRCC close to 1 or -1 indicates the corresponding parameters are influential.

## Results

To explore the effects of movement of human hosts between patches with different endemicities and MDAi control histories on mf prevalence and the ATP, we present the outcome of simulations with human movement originating in only one of the areas at a time (Fig 3).

**Figure 3.**
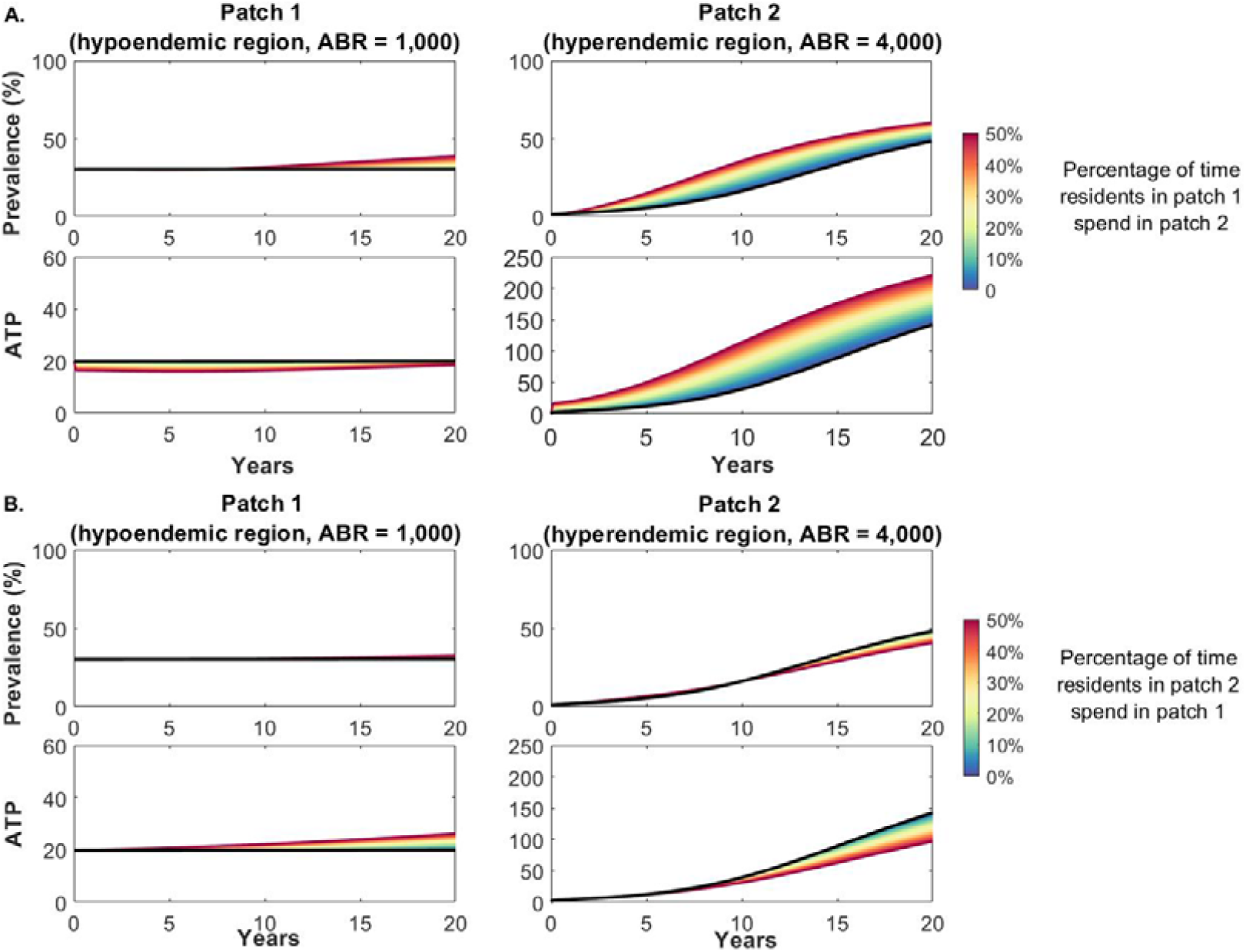
Change in microfilariae prevalence among population (aged ≥5) and annual transmission potential (ATP) in patch 1 and patch 2 due to human hosts aged 20 to 50 years (35% of the population) travelling between the patches for the first 20 years after MDAi was discontinued in patch 2. Note the difference in scale for the ATP of patch 1 and patch 2. **A**. Effect of individuals living in the hypoendemic area (patch 1) spending a percentage of their time in the post-MDAi hyperendemic area (patch 2). In this scenario the fraction of time residents in patch 1 spend visiting patch 2 (*f*_12_) is varied while assuming that residents of patch 2 do not travel to patch 1 (*f*_21_ = 0). **B**. Effect of individuals living in patch 2 spending a percentage of their time in the hypoendemic area (patch 1). In this scenario the parameter *f*_21_ is varied while assuming that residents of patch 1 do not travel to patch 2 *(f*_12_ = 0). Solid black lines indicate prevalence and ATP if the two patches are not coupled; i.e., when residents of one area do not spend time in the other. See Fig S1 in S1 File for results over the time scale required to reach equilibrium mf prevalence.

### Effect of human movement on mf prevalence and ATP in two connected areas

#### Effect on mf prevalence and ATP when individuals from the hypoendemic patch 1 visit the post-MDAi hyperendemic patch 2

Recrudescence in the post-MDAi hyperendemic area (patch 2) is accelerated by visiting residents of the untreated hypoendemic area (patch 1) travelling to the formerly hyperendemic area (patch 2) (Fig 3A). The infected 20- to 50-year-old visitors from patch 1 (initially ∼24.8% of these visitors) serve as a source of mf for vectors in patch 2, and these vectors in turn infect patch 2 residents. As infection prevalence among the residents of patch 2 increases, they (as well as their visitors from patch 1) serve as additional sources of mf for the vectors in patch 2, resulting in acceleration of the increase in mf prevalence and the ATP in patch 2. Moreover, as the ATP in patch 2 increases, mf prevalence in patch 1 also increases, because new infections acquired by patch 1 residents during their visits to patch 2 who then return to their “home” in patch 1 become more frequent. The initial decline in the ATP in the hypoendemic patch 1 arises from the system being in equilibrium at the start of the simulation. When human movement from patch 1 into patch 2 is introduced, the initial infected patch 1 residents who visit patch 2 are unavailable to patch 1 vectors for the duration of their visit and thus the ATP and prevalence in patch 1 decline transiently. Subsequently, as both mf prevalence and the ATP in patch 2 rise, patch 1 residents become infected while they are in patch 2, resulting in increasing mf prevalence among patch 1 residents, and thus increasing ATP in patch 1 (Fig 3A). Note that the pOTTIS does not necessarily represent the breakpoint below which transmission spontaneously decays to zero [40]. Thus, under the conditions being modeled in patch 2 (hyperendemic with high ABR and no vector control), the modelled prevalence will always rebound in patch 2 when MDAi is stopped. The important point illustrated in Fig 3 is that coupling of hyperendemic patch 2 with a hypoendemic patch 1 significantly *accelerates* this rebound over the 5 – 10 years following MDAi cessation in patch 2.

#### Effect on mf prevalence and ATP when individuals from the post-MDAi hyperendemic patch 2 visit the hypoendemic patch 1

When residents of the post-MDAi hyperendemic area (patch 2) visit the hypoendemic area (patch 1), the impact on mf prevalence among the residents of the hypoendemic area is more gradual and the magnitude of the change in mf prevalence is smaller when compared to the impact of human movement from patch 1 to patch 2 (Fig 3B). Mf from patch 2 residents (mf prevalence ∼1.1% in those aged 20-50 who travel to patch 1) act as an additional source of infection for the vectors in patch 1, which is evident in the slowly increasing ATP. However, mf prevalence only sees modest increases as the residents of patch 2 only spend part of their time in patch 1 and the patch 1 ABR is low. The more time residents of the post-MDAi hyperendemic area spend in the hypoendemic area (and thus are ‘available’ as source of infection of vectors in the hypoendemic area, but not in the post-MDAi hyperendemic area during that time), the earlier and faster the increase in the mf prevalence and ATP in patch 1 and the slower the increase in patch 2 (Fig 3B).

### Effects of the ABR in areas connected by human movement on mf prevalence

To consider the influence of the patch-specific ABR on mf prevalence over time, we selected values for movement parameters and varied the ABR in each patch.

#### Effect of variation of ABR in the hypoendemic area on mf prevalence

Fig 4 demonstrates the impact of different ABRs in the hypoendemic area (patch 1). The range of values for *ABR*_1_ were chosen to be representative of hypoendemic areas (Table S2 in S1_File). With an ABR of less than approximately 750, an isolated patch would not be able to sustain transmission (Fig 4A,C). Not surprisingly, the higher the ABR in patch 1 is, the faster resurgence in patch 2 occurs (Fig 4B,D). As already demonstrated in Fig 3, the rate of resurgence is affected by the direction of movement, with a faster rate of resurgence when residents of patch 1 spend time in patch 2 compared to movement in the opposite direction. Furthermore, the ABR in patch 1 has a higher impact on the rate of resurgence in patch 2 when patch 1 residents travel to patch 2 than vice versa (Fig 4B,D). Importantly, by connecting a hypoendemic area with an ABR below the threshold biting rate (TBR) required to sustain transmission in isolated hypoendemic areas (∼ 750, see below) to a post-MDAi hyperendemic area, we observe that transmission can be sustained in patch 1 (Fig 4A,C). The lowest patch 1 TBR and time to sustainable transmission is affected by the direction of human movement: movement from the hypoendemic area into the hyperendemic area results in achieving sustained transmission in patch 1 with a lower patch 1 TBR than movement in the other direction (Fig 4A,C; Fig S2 in S1 File).

**Figure 4.**
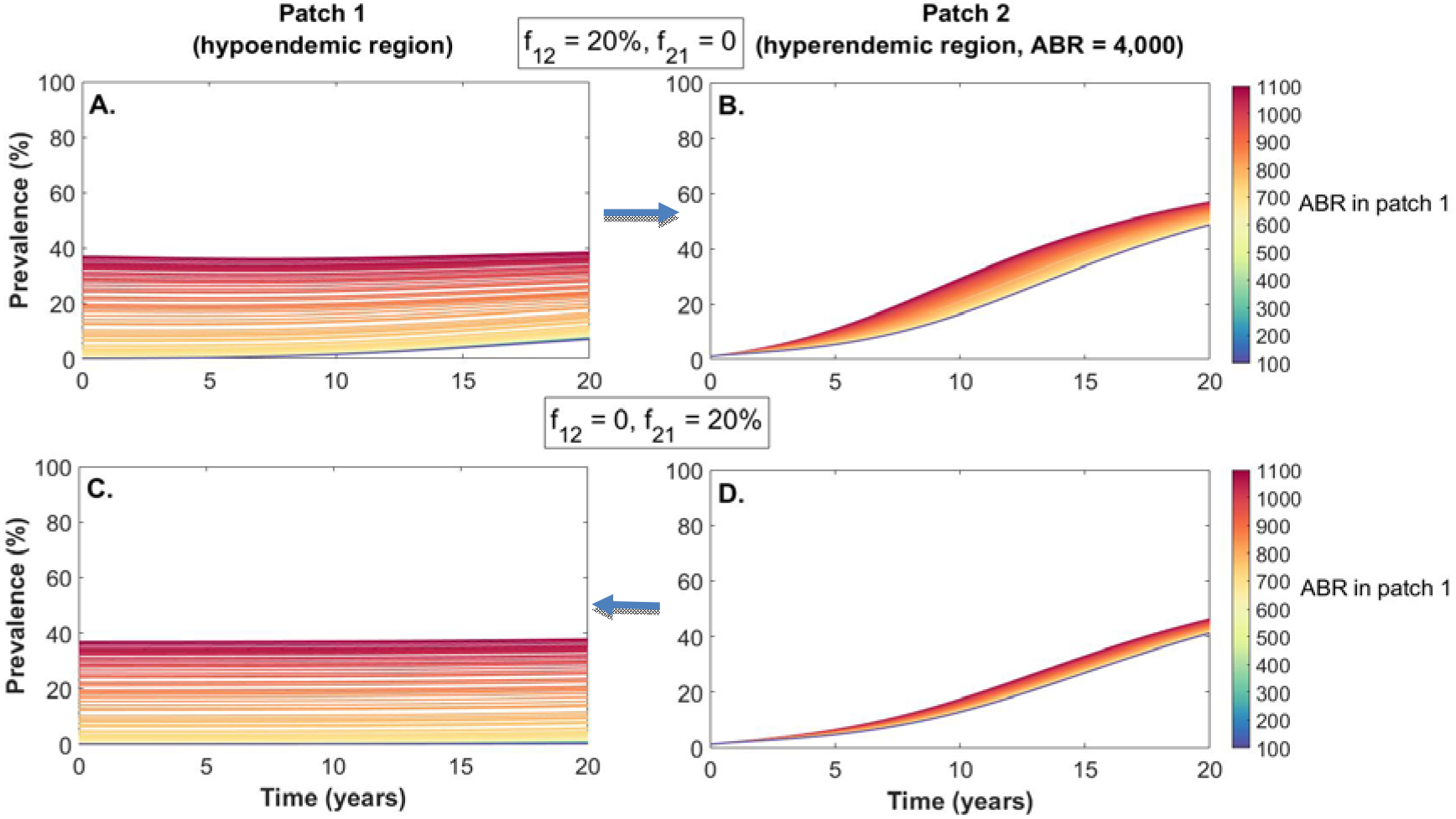
Impact of variation in the Annual Biting Rate (ABR) of blackfly vectors in a hypoendemic area (patch 1) on microfilariae prevalence (in ages 5 and above) when connected through human movement to a hyperendemic area (patch 2) for the first 20 years after MDAi was discontinued in patch 2. Patch 1 is initialized by running the model assuming patch 1 is isolated (i.e. not connected to patch 2 through human movement) to equilibrium for each ABR_1_ value; Patch 2 is initialized with <1.4% mf prevalence across all ages. See Fig S2 in S1 File for results over the time scale required to reach equilibrium prevalence.

#### Effect of variation of ABR in the hyperendemic area on mf prevalence

Fig 5 demonstrates the impact of different ABRs in the post-MDAi hyperendemic area (patch 2). The range of values for *ABR*_2_ were chosen to be representative of a hyperendemic area (Table S2 in S1_File) which correspond to mf prevalences between 61% and 82% in the population aged 5 years and above for an isolated patch without interventions. Not surprisingly, the higher the ABR in patch 2 is, the faster resurgence in patch 2 occurs. The rate of resurgence is only marginally affected by the direction of movement (Fig 4B,D). In contrast, the degree to which mf prevalence in the hypoendemic area (patch 1) increases depends on the direction of movement, with movement from patch 1 to patch 2 having a larger impact on the mf prevalence in the hypoendemic area (patch 1) than movement from patch 2 to patch 1 (Fig 4A,C).

**Figure 5.**
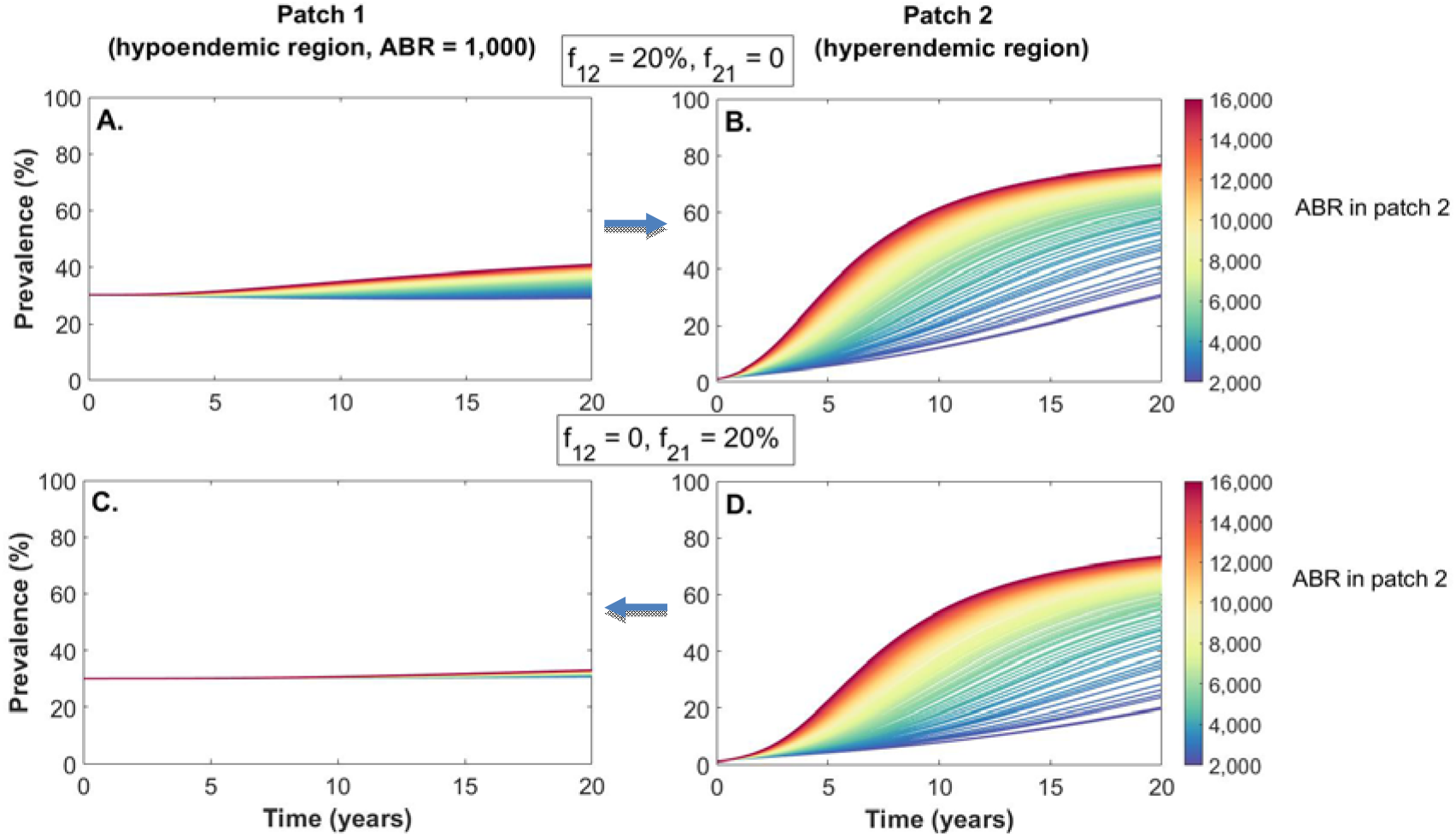
Impact variation in the ABR in a hyperendemic area (patch 2) which discontinued MDAi has on mf prevalence (in ages 5 and above) when connected through human movement to a hypoendemic area (patch 1) during the 20 years post-MDAi. Patch 1 is initialized by running the model assuming patch 1 is not connected to patch 2 to equilibrium (with ABR_1_= 1,000); Patch 2 is initialized with <1.4% mf prevalence across all ages. See Fig S3 in S1 File for results over the time scale required to reaching equilibrium in mf prevalence.

### Effect of human movement on local transmission in a hypoendemic area

For the set of parameters provided in Table S2 in A1_File, parasite transmission cannot be sustained in an isolated patch with an ABR of less than ≈ 750. In the presence of human movement between a hypoendemic patch and a hyperendemic patch, local transmission in the hypoendemic patch is possible at lower ABRs (Fig S5 in S1_File).

### Sensitivity of modeled long-term mf prevalence to variation in factors impacting mf prevalence

The ranking of the most influential parameters impacting long-term mf prevalence emerging from the Partial Rank Correlation Coefficient (PRCC) method differed between the hypo- and hyperendemic areas (Fig 6). The top three parameters that have a strong positive association with prevalence in both patches were the patch-specific ABR, the proportion of L3 larvae developing to adult worms within the human host when ATP *→* 0 (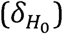), and the proportion of blood meals vectors take on humans relative to other mammals (the human blood index, *h*) (see Table S2 in S1_File for further details). Not surprisingly, the mortality rate of microfilariae within the human host (*σ*_*M*_) had the strongest negative association with long-term mf prevalence in both areas, indicating a decrease in mf prevalence with increase in mf mortality rate. The parameter quantifying movement from patch 1 to patch 2 (*f*_12_) has a moderate positive association with mf prevalence in patch 1 (Fig 3A), whereas movement from patch 2 to patch 1 (*f*_21_) has a moderate negative association with mf prevalence in patch 2 (Fig 3B).

**Figure 6.**
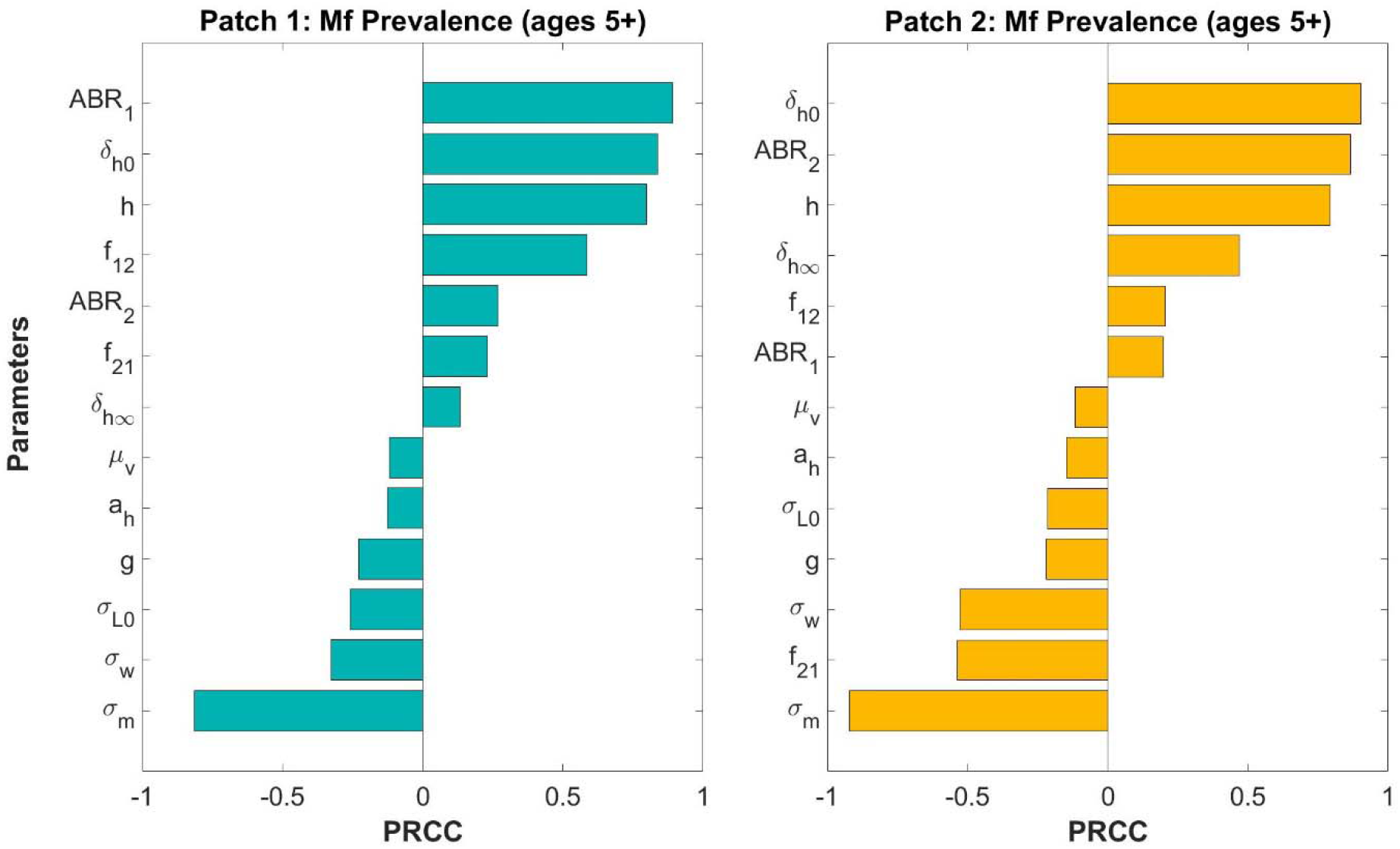
Tornado plots for partial rank correlation coefficient (PRCC) results from 1,000 samples using Latin hypercube sampling (LHS) illustrating drivers of long-term mf prevalence of population aged 5+ in each patch. Only statistically significant parameters are shown (evaluated at α = 0.01). PRCC values close to 1 or -1 indicate most influential parameters. Because the parameters are ranked by the strength of the association, the order of parameters is different for each patch. For explanation of parameters, see Table S2 in S1 File.

One factor impacting TBR is the human blood index, the proportion of blood meals the vector takes on humans (see Table S2 in S1 File). Thus, we conducted a one-way sensitivity analysis to determine the impact of the human blood index on the relationship between long-term mf prevalence and ABR in an isolated population. Through a simulation study, the TBR for a single isolated patch with input parameters as defined in Table S2 in S1_File is estimated to be approximately 750. However, if the human blood index is increased from 0.3 (as estimated in Cameroon [87]) to 0.67 (as estimated in Burkina Faso and Côte d’Ivoire [88]), the TBR decreases significantly (Fig S6 in S1_File). The human blood index is likely to vary in different onchocerciasis-endemic settings, because it is dependent both on the vector species present and their anthropophily and anthropophagy and on the availability and density of domesticated and wild species including chicken, goats, or cattle [87, 89-93].

## Discussion

We have adapted the EPIONCHO framework described by Filipe et al. [46] to produce a patch model for evaluating the effect of human and vector movement between any number of areas (patches) that differ with respect to key factors important for onchocerciasis elimination efforts, including ABR, MDAi history, and mf prevalence. We have demonstrated the type of information this patch model can provide by modeling a simple scenario: the impact of unidirectional travel of people between two areas with different endemicity and MDAi history on changes in mf prevalence and ATP in each of these areas.

The results illustrate that the risk of recrudescence in a hyperendemic area post-MDAi (patch 2) is increased, and recrudescence accelerated, when there is human movement between that area and a hypoendemic area without ongoing interventions (patch 1) (Fig 3, Fig S1 in S1_File). We have also shown that movement of humans between a hyperendemic area (patch 2) and a hypoendemic area (patch 1) can enable sustained transmission in the hypoendemic area at ABRs which would not support local transmission in an isolated area (Fig 4, Fig S2 in S1_File, Fig S5 in S1_File). This finding is consistent with the conclusions of a recent modeling study utilizing the stochastic, individual-based framework ONCHOSIM to examine *O. volvulus* transmission between connected villages [56]. The results presented here further indicate that the direction and extent of human movement between areas cause variation in the strength and direction (positive or negative association) of the effect key parameters (in particular, the ABR) have on the long-term mf prevalence in areas with different endemicity and initial mf prevalence.

It is important to note the time frame over which changes in mf prevalence and ATP are predicted to occur. In the hyperendemic area, they remain low until nearly 10 years post-MDAi (Fig 3B,D) and will be challenging to detect with available methods during the (on average) 3-5 years of post-MDAi surveillance currently recommended for confirming interruption of parasite transmission [51]. In our model, the rate of increase in mf prevalence and ATP is determined by the degree and direction of movement from and to the hypoendemic patch (coupling) as well as by the ABRs in both patches. Greater coupling and higher ABRs result in faster resurgence of mf prevalence and ATP. These dependencies imply that the duration of post-MDAi surveillance to confirmation of interruption of parasite transmission need to be chosen in view of the distribution of ABRs across the whole transmission zone, i.e. the whole area over which parasites are transmitted through human (and vector) movement. Under a scenario where MDAi brings the prevalence to zero in the hyperendemic region, or where there is stochastic loss of infection, then this predicted return to prevalence would presumably take longer. The scenarios explored here assumed that the starting mf prevalence in the post-MDAi hyperendemic patch 2 was below the epidemiological pOTTIS (adapted for modelling to <1.4% mf prevalence across all ages). As outlined above, pOTTIS is defined as “provisional” [31], and neither the mf prevalence-based nor the vector infectivity-based pOTTIS in the WHO 2016 guidelines represent parasite transmission breakpoints, which have been described as ‘hypothetical and [of] elusive nature’ [40]. Our patch model predicts that an isolated patch that has an ABR high enough to sustain transmission will experience resurgence to pre-intervention levels of infection after cessation of MDAi (see black line for patch 2 presented in Fig 3 and Fig S1 in S1 File, and as previously described [40]).

Due to the deterministic nature of the model, there is no potential for stochastic fade-out of infection in a patch even at very low prevalence. Accordingly, should prevalence in a hyperendemic area approach zero, our patch model might predict an earlier resurgence than may be observed in a real-world scenario. However, the choice to use a deterministic model does not impact the model’s findings on the key drivers of the rate of recrudescence when coupling between areas is incorporated.

We confirmed that the relationship between ABR and modeled long-term mf prevalence and ATP in an area depends on the input parameters, which in turn depend on the context being modeled (S1 Appendix Fig S5). We found that the ABR, the proportion of L3 larvae developing to adult worms within the human host when ATP → 0 (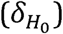) and the human blood index (*h*) are key drivers (most influential parameters) of the modeled long-term mf prevalence in both the hypoendemic and hyperendemic area (Fig 6, S1 Appendix Table S1). While these are not novel findings, it is important to recall that parameterizing a model for different contexts will lead to different outputs including the TBR, estimates of time to reach specified thresholds with specified interventions [39, 71] or, in our case, time course of changes in mf prevalence and ATP in the absence of interventions (Fig. 4, Fig. 5)). Our results suggest that the extent of parasite transmission between areas within a transmission zone differing in key input parameter characteristics through human (and by extension vector) movement should also be included in the models. The challenge, then, is to collect the data that are required to estimate these input parameters in a range of epidemiological and ecological settings or, ideally, specific geographic areas of interest. Michael and colleagues [49] proposed a local, data-driven approach to model parameterization using available baseline pre-intervention and follow-up mf prevalence data, area-wide pre-intervention ABR, and information on interventions from the hyperendemic areas Itwara and Kashoya Kitomi and the mesoendemic Bwindi area in Western Uganda.

To better parameterize the coupling strength in this model, we require good estimates of human population mobility, which can be derived by making use of more sophisticated population mobility models (such as the gravity or radiation models or a combination of these [94]. Furthermore, getting a better understanding of the impact that frequency and duration of human mobility between areas has on the probability of parasite transmission between areas is a critical next step. A high number of trips between areas with short duration may not have the same contribution to transmission dynamics in the destination area as a low number of trips with long duration and the impact of travel is likely to depend on the timing relative to transmission season. This requires adapting the current model to allow for a dynamic human population or adaptation of an individual-based model (EPIONCHO-IBM [47] or ONCHOSIM [56]) to take such characteristics into account.

While vector control is not a parameter specified in the model, the effect of vector control (e.g., via larviciding [95, 96] or community-directed slash-and-clear [97-99] is to reduce ABR. The ABR determines the likelihood that, should MDAi be stopped, a population of parasites that had reached the epidemiology-based or entomology-based pOTTIS would subsequently rebound, such that recrudescence will not occur when ABR is sufficiently low [40]. We compared the effect of varying patch-specific ABR on transmission in both patches (Fig 5). Our results suggest that vector control that reduces ABR in the hyperendemic patch 2 could significantly slow the rate of recrudescence due to travel from an untreated hypoendemic patch 1 into the post-MDAi patch 2. Reducing ABR in the post-MDAi patch 2 may also reduce mf prevalence in the hypoendemic patch 1, and would have a larger impact than vector control in patch 1 alone (Fig. 4). Thus, we are working towards incorporating data-based estimates of vector migration and on incorporating temporal variation in vector abundance due to interventions.

We have focused here on how spatial heterogeneity of pre-control prevalence might impact post-MDAi prevalence and recrudescence, and find that recrudescence can be driven by movement from untreated hypoendemic regions. Temporal heterogeneity in the frequency and duration of MDAi is also likely to impact post-MDAi prevalence in connected areas and by extension the risk of recrudescence. Temporal heterogeneity in MDAi has been explored in an individual-based, stochastic framework [56]. Incorporating temporal heterogeneity into the spatial, deterministic, population-based model described here to explore whether the predicted outcomes for spatial and temporal heterogeneity are similar would strengthen the evidence for making decisions about MDAi timetables (see [40]).

## Conclusions

Our study highlights the need to consider imported infection through mobility of human or vector hosts between connected regions when the goal is elimination of transmission in the vast, contiguous, and heterogeneous areas of sub-Saharan Africa. When determining the geographic scale at which stop-MDAi decisions should be made, treating any given location as though it were isolated from the surrounding region is unrealistic and may jeopardize elimination efforts [100].

We chose the scenarios modeled in view of two questions that need to be answered to achieve sustained and cost-effective elimination of *O. volvulus* transmission: 1) what are the characteristics of hypoendemic areas not previously included in MDAi where interventions need to be implemented, and 2) what are the geographic areas that belong to a transmission zone and that need to be included in evaluations for decisions on stopping MDAi and conducting post-treatment and post-elimination surveillance [51]. Our results, along with decades of epidemiological and entomological data from programs that have demonstrated transmission of onchocerciasis among communities, suggest that the answers to both questions need to consider not only the ABRs but also the transport of parasites via human movement. The key missing parameter is an evidence-based understanding of the scale over which transmission connectivity extends, and a method by which that transmission connectivity can be measured [3]. This emphasizes not only the importance of decisions on which hypoendemic areas to include in MDAi (with or without complementary control measures or alternative treatment strategies), but also implies that the boundaries of transmission zones are unlikely to correspond to the administrative boundaries within which responsibility for interventions may be with different health system units (i.e., “implementation units” such as “APOC project areas”) both within and across countries. This needs to be considered when planning (a) the geographic areas and the density of sampling for evaluations for decisions on when and where to stop MDAi, (b) where, how frequently and for how long to conduct post-MDAi surveillance for verification of elimination of transmission and (c) where and how frequently to conduct post-elimination surveillance. Until suitable methods for delineation of transmission zones are available [3, 101], a pragmatic approach is to assume parasite transmission is very likely between endemic areas within locally typical day-to-day travelling distances (and within vector flight distance).

Consideration of cost-effectiveness includes (a) past investments into onchocerciasis control and elimination, (b) investments still needed to achieve sustainable elimination, and (c) other public health objectives ‘competing’ for the same funds. The timeframe of the modelled increase in mf prevalence and ATP in the post MDAi area deserves consideration in decisions on the risk of and timeframe of resurgence acceptable to stakeholders. Given the cost implications (including opportunity cost) of stopping interventions too early or later than needed, we encourage the development and application of models, such as the one presented here, that explicitly consider human and vector movement in a heterogeneous landscape (see also [56]) and that can be applied to explore the appropriate geographic scale for decision making about the location and duration of MDAi. Such models, ideally parameterized for specific geographic areas, can provide national program managers with a better estimate of the risk of and time course of recrudescence when interventions are stopped too early, or in only parts of a transmission zone when there is ongoing transmission elsewhere, and serve as input into relevant economic analysis.

## Data Availability

The code for the model is available through GitHub.

https://github.com/shedtke/Onchocerciasis_patch_model

## Acknowledgments

The authors wish to acknowledge Wilma Stolk, Luc Coffeng, Rebecca Chisholm, Shlomo Riesenberg Xu, and Himal Shrestha for helpful discussions.

The authors alone are responsible for the views expressed which do not necessarily represent the views, decisions, or policies of the institutions with which the authors are affiliated.

## Copyright

2022 World Health Organization. This is an open access article distributed under the Creative Commons Attribution IGO License, which permits unrestricted use, distribution, and reproduction in any medium, provided the original work is properly cited. https://creativecommons.org/licenses/by/3.0/igo/. In any use of this article, there should be no suggestion that WHO endorses any specific organization, products or services. The use of the WHO logo is not permitted. This notice should be preserved along with the article’s original URL.

## Supporting information Legend

S1 File: Model equations, parameter table, and supplemental figures.

## References

1. Remme JHF, Boatin B, Boussinesq M. Helminthic diseases: Onchocerciasis and loiasis. In: Quah SR, Cockerham W, editors. International Encyclopedia of Public Health. Second ed: Elsevier Inc.; 2017. p. 576–87.

2. World Health Organization. Elimination of human onchocerciasis: progress report, 2018-2019. Weekly Epidemiological Record. 2019;94:513–24.

3. Hedtke SM, Kuesel AC, Crawford KE, Graves PM, Boussinesq M, Lau CL, et al. Genomic epidemiology in filarial nematodes: transforming the basis for elimination program decisions. Front Genet. 2020;10:1282. Epub 2020/01/31. doi: 10.3389/fgene.2019.01282. PubMed PMID: 31998356; PubMed Central PMCID: PMCPMC6964045.

4. World Health Organization. Elimination of human onchocerciasis: progress report, 2020. Weekly Epidemiological Record. 2021;46(96):557–67.

5. Dadzie Y, Neira M, Hopkins D. Final report of the conference on the eradicability of onchocerciasis. Filaria J. 2003;2:2.

6. Diawara L, Traore MO, Badji A, Bissan Y, Doumbia K, Goita SF, et al. Feasibility of onchocerciasis elimination with ivermectin treatment in endemic foci in Africa: first evidence from studies in Mali and Senegal. PLoS Negl Trop Dis. 2009;3(7):e497. doi: 10.1371/journal.pntd.0000497. PubMed PMID: 19621091; PubMed Central PMCID: PMCPMC2710500.

7. Tekle AH, Elhassan E, Isiyaku S, Amazigo UV, Bush S, Noma M, et al. Impact of long-term treatment of onchocerciasis with ivermectin in Kaduna State, Nigeria: first evidence of the potential for elimination in the operational area of the African Programme for Onchocerciasis Control. Parasit Vectors. 2012;5:28. doi: 10.1186/1756-3305-5-28. PubMed PMID: 22313631; PubMed Central PMCID: PMCPMC3296569.

8. Traore MO, Sarr MD, Badji A, Bissan Y, Diawara L, Doumbia K, et al. Proof-of-principle of onchocerciasis elimination with ivermectin treatment in endemic foci in Africa: final results of a study in Mali and Senegal. PLoS Negl Trop Dis. 2012;6(9):e1825. doi: 10.1371/journal.pntd.0001825. PubMed PMID: 23029586; PubMed Central PMCID: PMCPMC3441490.

9. Tekle AH, Zoure HG, Noma M, Boussinesq M, Coffeng LE, Stolk WA, et al. Progress towards onchocerciasis elimination in the participating countries of the African Programme for Onchocerciasis Control: epidemiological evaluation results. Infect Dis Poverty. 2016;5(1):66. doi: 10.1186/s40249-016-0160-7. PubMed PMID: 27349645; PubMed Central PMCID: PMCPMC4924267.

10. World Health Organization, African Programme for Onchocerciasis Control. Eighteenth session of the Join Action Forum, Bujumbura, Burundi, 11-13 Dec 2021: final communiqué. Geneva: 2012 Contract No.: JAF18.

11. World Health Organization. Ending the neglect to attain the sustainable development goals: a road map for neglected tropical diseases 2021–2030. Geneva: 2020 2020. Report No.: Contract No.: WHO/UCN/NTD/2020.01.

12. Turner HC, Walker M, Attah SK, Opoku NO, Awadzi K, Kuesel AC, et al. The potential impact of moxidectin on onchocerciasis elimination in Africa: an economic evaluation based on the Phase II clinical trial data. Parasit Vectors. 2015;8:167. doi: 10.1186/s13071-015-0779-4. PubMed PMID: 25889256; PubMed Central PMCID: PMCPMC4381491.

13. World Health Organization, African Programme for Onchocerciasis Control. Report of the consultative meetings on strategic options and alternative treatment strategies for accelerating onchocerciasis elimination in Africa. 2015 WHO/MG/15.20.

14. Al-Kubati AS, Mackenzie CD, Boakye D, Al-Qubati Y, Al-Samie AR, Awad IE, et al. Onchocerciasis in Yemen: moving forward towards an elimination program. Int Health. 2018;10(Suppl_1):i89–i96. doi: 10.1093/inthealth/ihx055. PubMed PMID: 29471343.

15. Boakye D, Tallant J, Adjami A, Moussa S, Tekle A, Robalo M, et al. Refocusing vector assessment towards the elimination of onchocerciasis from Africa: a review of the current status in selected countries. Int Health. 2018;10(Suppl_1):i27–i32. doi: 10.1093/inthealth/ihx066. PubMed PMID: 29471346; PubMed Central PMCID: PMCPMC5881273.

16. Cantey PT, Roy SL, Boakye D, Mwingira U, Ottesen EA, Hopkins AD, et al. Transitioning from river blindness control to elimination: steps toward stopping treatment. Int Health. 2018;10(suppl_1):i7–i13. doi: 10.1093/inthealth/ihx049. PubMed PMID: 29471338; PubMed Central PMCID: PMCPMC5881257.

17. Elhassan E, Zhang Y, Bush S, Molyneux D, Kollmann MKH, Sodahlon Y, et al. The role of the NGDO Coordination Group for the Elimination of Onchocerciasis. Int Health. 2018;10(suppl_1):i97–i101. Epub 2018/02/23. doi: 10.1093/inthealth/ihx050. PubMed PMID: 29471339.

18. Griswold E, Unnasch T, Eberhard M, Nwoke BEB, Morales Z, Muheki Tukahebwa E, et al. The role of national committees in eliminating onchocerciasis. Int Health. 2018;10(suppl_1):i60–i70. Epub 2018/02/23. doi: 10.1093/inthealth/ihx048. PubMed PMID: 29471337.

19. Katabarwa MN, Lakwo T, Habomugisha P, Unnasch TR, Garms R, Hudson-Davis L, et al. After 70 years of fighting an age-old scourge, onchocerciasis in Uganda, the end is in sight. Int Health. 2018;10(suppl_1):i79–i88. Epub 2018/02/23. doi: 10.1093/inthealth/ihx044. PubMed PMID: 29471335.

20. Opoku NO, Bakajika DK, Kanza EM, Howard H, Mambandu GL, Nyathirombo A, et al. Single dose moxidectin versus ivermectin for Onchocerca volvulus infection in Ghana, Liberia, and the Democratic Republic of the Congo: a randomised, controlled, double-blind phase 3 trial. Lancet. 2018;392(10154):1207–16. doi: 10.1016/S0140-6736(17)32844-1. PubMed PMID: 29361335; PubMed Central PMCID: PMCPMC6172290.

21. Rebollo MP, Zoure H, Ogoussan K, Sodahlon Y, Ottesen EA, Cantey PT. Onchocerciasis: shifting the target from control to elimination requires a new first-step-elimination mapping. Int Health. 2018;10(suppl_1):i14–i9. doi: 10.1093/inthealth/ihx052. PubMed PMID: 29471341.

22. Unnasch TR, Golden A, Cama V, Cantey PT. Diagnostics for onchocerciasis in the era of elimination. Int Health. 2018;10(suppl_1):i20–i6. doi: 10.1093/inthealth/ihx047. PubMed PMID: 29471336.

23. Boussinesq M, Fobi G, Kuesel AC. Alternative treatment strategies to accelerate the elimination of onchocerciasis. Int Health. 2018;10(suppl_1):i40–i8. doi: 10.1093/inthealth/ihx054. PubMed PMID: 29471342.

24. Zouré HG, Noma M, Tekle AH, Amazigo UV, Diggle PJ, Giorgi E, et al. The geographic distribution of onchocerciasis in the 20 participating countries of the African Programme for Onchocerciasis Control: (2) pre-control endemicity levels and estimated number infected. Parasit Vectors. 2014;7:326. doi: 10.1186/1756-3305-7-326. PubMed PMID: 25053392; PubMed Central PMCID: PMCPMC4222889.

25. O’Hanlon SJ, Slater HC, Cheke RA, Boatin BA, Coffeng LE, Pion SD, et al. Model-based geostatistical mapping of the prevalence of Onchocerca volvulus in West Africa. PLoS Negl Trop Dis. 2016;10(1):e0004328. Epub 2016/01/16. doi: 10.1371/journal.pntd.0004328. PubMed PMID: 26771545; PubMed Central PMCID: PMCPMC4714852.

26. Schmidt CA, Cromwell EA, Hill E, Donkers KM, Schipp MF, Johnson KB, et al. The prevalence of onchocerciasis in Africa and Yemen, 2000-2018: a geospatial analysis. BMC Med. 2022;20(1):293. Epub 2022/09/07. doi: 10.1186/s12916-022-02486-y. PubMed PMID: 36068517; PubMed Central PMCID: PMCPMC9449300.

27. UNDP/World Bank/WHO Special Programme for Research and Training in Tropical Diseases. Methods for community diagnosis of onchocerciasis to guide ivermectin based control in Africa: report of an informal consultation held in Ouagadougou from 19-21 November 1991. Geneva: World Health Organization; 1992.

28. Ngoumou P, Walsh JF, WHO Programme for the Prevention of Blindness, UNDP/World Bank/WHO Special Programme for Research and Training in Tropical Diseases. A manual for rapid epidemiological mapping of onchocerciasis. 1993.

29. Noma M, Nwoke BE, Nutall I, Tambala PA, Enyong P, Namsenmo A, et al. Rapid epidemiological mapping of onchocerciasis (REMO): its application by the African Programme for Onchocerciasis Control (APOC). Ann Trop Med Parasitol. 2002;96 Suppl 1:S29–39. PubMed PMID: 12081248.

30. Noma M, Zouré HG, Tekle AH, Enyong PA, Nwoke BE, Remme JH. The geographic distribution of onchocerciasis in the 20 participating countries of the African Programme for Onchocerciasis Control: (1) priority areas for ivermectin treatment. Parasit Vectors. 2014;7:325. doi: 10.1186/1756-3305-7-325. PubMed PMID: 25053266; PubMed Central PMCID: PMCPMC4223657.

31. World Health Organization, African Programme for Onchoceriasis Control. Conceptual and operational framework of onchocerciasis elimination with ivermectin treatment. In: APOC/WHO, editor. Burkina Faso: World Health Organization; 2010.

32. World Health Organization, African Programme for Onchocerciasis Control. Guidelines for revising ivermectin treatment boundaries within the context of onchocerciasis elimination. 2015.

33. de Vos AS, Stolk WA, de Vlas SJ, Coffeng LE. The effect of assortative mixing on stability of low helminth transmission levels and on the impact of mass drug administration: Model explorations for onchocerciasis. PLoS Negl Trop Dis. 2018;12(10):e0006624. Epub 2018/10/09. doi: 10.1371/journal.pntd.0006624. PubMed PMID: 30296264; PubMed Central PMCID: PMCPMC6175282.

34. Plaisier AP, van Oortmarssen GJ, Habbema JD, Remme J, Alley ES. ONCHOSIM: a model and computer simulation program for the transmission and control of onchocerciasis. Comput Methods Programs Biomed. 1990;31(1):43–56. PubMed PMID: 2311368.

35. Remme JHF, Alley ES, Plaisier AP. Estimation and prediction in tropical disease control: the example of onchocerciasis. In: Mollison D, editor. Epidemic models: their structure and relation to data. Cambridge: Cambridge University Press; 1995. p. 372–92.

36. Winnen M, Plaisier AP, Alley ES, Nagelkerke NJ, van Oortmarssen G, Boatin BA, et al. Can ivermectin mass treatments eliminate onchocerciasis in Africa? Bull World Health Organ. 2002;80(5):384–91.

37. Turner HC, Churcher TS, Walker M, Osei-Atweneboana MY, Prichard RK, Basáñez MG. Uncertainty surrounding projections of the long-term impact of ivermectin treatment on human onchocerciasis. PLoS Negl Trop Dis. 2013;7(4):e2169. doi: 10.1371/journal.pntd.0002169. PubMed PMID: 23634234; PubMed Central PMCID: PMCPMC3636241.

38. Coffeng LE, Stolk WA, Hoerauf A, Habbema D, Bakker R, Hopkins AD, et al. Elimination of African onchocerciasis: modeling the impact of increasing the frequency of ivermectin mass treatment. PLoS One. 2014;9(12):e115886. doi: 10.1371/journal.pone.0115886. PubMed PMID: 25545677; PubMed Central PMCID: PMCPMC4278850.

39. Stolk WA, Walker M, Coffeng LE, Basáñez Mg, de Vlas SJ. Required duration of mass ivermectin treatment for onchocerciasis elimination in Africa: a comparative modelling analysis. Parasit Vectors. 2015;8:552. doi: 10.1186/s13071-015-1159-9. PubMed PMID: 26489937; PubMed Central PMCID: PMCPMC4618738.

40. Basáñez Mg, Walker M, Turner HC, Coffeng LE, de Vlas SJ, Stolk WA. River blindness: mathematical models for control and elimination. Adv Parasitol. 2016;94:247–341. doi: 10.1016/bs.apar.2016.08.003. PubMed PMID: 27756456.

41. Behrend MR, Basáñez Mg, Hamley JID, Porco TC, Stolk WA, Walker M, et al. Modelling for policy: The five principles of the Neglected Tropical Diseases Modelling Consortium. PLoS Negl Trop Dis. 2020;14(4):e0008033. Epub 2020/04/10. doi: 10.1371/journal.pntd.0008033. PubMed PMID: 32271755; PubMed Central PMCID: PMCPMC7144973 following competing interests: MRB is a contractor to the Bill & Melinda Gates Foundation. The other authors declare no competing interests.

42. World Health Organization, African Programme for Onchocerciasis Control. Report of the CSA advisory group on onchocerciasis elimination. 2011.

43. Dyson L, Stolk WA, Farrell SH, Hollingsworth TD. Measuring and modelling the effects of systematic non-adherence to mass drug administration. Epidemics. 2017;18:56–66. Epub 2017/03/11. doi: 10.1016/j.epidem.2017.02.002. PubMed PMID: 28279457; PubMed Central PMCID: PMCPMC5340860.

44. Basáñez Mg, Boussinesq M. Population biology of human onchocerciasis. Philos Trans R Soc Lond B Biol Sci. 1999;354(1384):809–26. doi: 10.1098/rstb.1999.0433. PubMed PMID: 10365406; PubMed Central PMCID: PMCPMC1692549.

45. Basáñez Mg, Ricardez-Esquinca J. Models for the population biology and control of human onchocerciasis. Trends Parasitol. 2001;17(9):430–8. PubMed PMID: 11530355. 46.

46. Filipe JA, Boussinesq M, Renz A, Collins RC, Vivas-Martinez S, Grillet ME, et al. Human infection patterns and heterogeneous exposure in river blindness. Proc Natl Acad Sci U S A. 2005;102(42):15265–70. Epub 2005/10/12. doi: 10.1073/pnas.0502659102. PubMed PMID: 16217028; PubMed Central PMCID: PMCPMC1257694.

47. Hamley JID, Milton P, Walker M, Basáñez Mg. Modelling exposure heterogeneity and density dependence in onchocerciasis using a novel individual-based transmission model, EPIONCHO-IBM: Implications for elimination and data needs. PLoS Negl Trop Dis. 2019;13(12):e0007557. Epub 2019/12/06. doi: 10.1371/journal.pntd.0007557. PubMed PMID: 31805049; PubMed Central PMCID: PMCPMC7006940.

48. Michael E, Smith ME, Katabarwa MN, Byamukama E, Griswold E, Habomugisha P, et al. Substantiating freedom from parasitic infection by combining transmission model predictions with disease surveys. Nat Commun. 2018;9(1):4324. Epub 2018/10/20. doi: 10.1038/s41467-018-06657-5. PubMed PMID: 30337529; PubMed Central PMCID: PMCPMC6193962.

49. Michael E, Smith ME, Singh BK, Katabarwa MN, Byamukama E, Habomugisha P, et al. Data-driven modelling and spatial complexity supports heterogeneity-based integrative management for eliminating Simulium neavei-transmitted river blindness. Sci Rep. 2020;10(1):4235. Epub 2020/03/08. doi: 10.1038/s41598-020-61194-w. PubMed PMID: 32144362; PubMed Central PMCID: PMCPMC7060237.

50. Hamley JID, Walker M, Coffeng LE, Milton P, de Vlas SJ, Stolk WA, et al. Structural uncertainty in onchocerciasis transmission models influences the estimation of elimination thresholds and selection of age groups for seromonitoring. J Infect Dis. 2020;221(Suppl 5):S510–S8. Epub 2020/03/17. doi: 10.1093/infdis/jiz674. PubMed PMID: 32173745; PubMed Central PMCID: PMCPMC7289547.

51. World Health Organization. Guidelines for stopping mass drug administration and verifying elimination of human onchocerciasis: criteria and procedures. Geneva: World Health Organization; 2016.

52. Lont YL, Coffeng LE, de Vlas SJ, Golden A, de Los Santos T, Domingo GJ, et al. Modelling anti-Ov16 IgG4 antibody prevalence as an indicator for evaluation and decision making in onchocerciasis elimination programmes. PLoS Negl Trop Dis. 2017;11(1):e0005314. doi: 10.1371/journal.pntd.0005314. PubMed PMID: 28114304; PubMed Central PMCID: PMCPMC5289624.

53. Verver S, Walker M, Kim YE, Fobi G, Tekle AH, Zoure HGM, et al. How can onchocerciasis elimination in Africa be accelerated? Modeling the impact of increased ivermectin treatment frequency and complementary vector control. Clin Infect Dis. 2018;66(Suppl_4):S267–S74. doi: 10.1093/cid/cix1137. PubMed PMID: 29860291; PubMed Central PMCID: PMCPMC5982715.

54. Coffeng LE, Stolk WA, Golden A, de Los Santos T, Domingo GJ, de Vlas SJ. Predictive value of Ov16 antibody prevalence in different subpopulations for elimination of African onchocerciasis. Am J Epidemiol. 2019;188(9):1723–32. Epub 2019/05/08. doi: 10.1093/aje/kwz109. PubMed PMID: 31062838; PubMed Central PMCID: PMCPMC6735885.

55. Blok DJ, Kamgno J, Pion SD, Nana-Djeunga HC, Niamsi-Emalio Y, Chesnais CB, et al. Feasibility of onchocerciasis elimination using a “test-and-not-treat” strategy in Loa loa co-endemic Areas. Clin Infect Dis. 2021;72(12):e1047–e55. Epub 2020/12/09. doi: 10.1093/cid/ciaa1829. PubMed PMID: 33289025; PubMed Central PMCID: PMCPMC8204788.

56. de Vos AS, Stolk WA, Coffeng LE, de Vlas SJ. The impact of mass drug administration expansion to low onchocerciasis prevalence settings in case of connected villages. PLoS Negl Trop Dis. 2021;15(5):e0009011. Epub 2021/05/13. doi: 10.1371/journal.pntd.0009011. PubMed PMID: 33979331; PubMed Central PMCID: PMCPMC8143415.

57. Koala L, Nikiema A, Post RJ, Paré AB, Kafando CM, Drabo F, et al. Recrudescence of onchocerciasis in the Comoé valley in Southwest Burkina Faso. Acta Trop. 2017;166:96–105. doi: 10.1016/j.actatropica.2016.11.003. PubMed PMID: 27845063.

58. Nikièma AS, Koala L, Post RJ, Paré AB, Kafando CM, Drabo F, et al. Onchocerciasis prevalence, human migration and risks for onchocerciasis elimination in the Upper Mouhoun, Nakambe and Nazinon river basins in Burkina Faso. Acta Trop. 2018;185:176–82. doi: 10.1016/j.actatropica.2018.05.013. PubMed PMID: 29782820.

59. Koala L, Nikiema AS, Pare AB, Drabo F, Toe LD, Belem AMG, et al. Entomological assessment of the transmission following recrudescence of onchocerciasis in the Comoé Valley, Burkina Faso. Parasit Vectors. 2019;12(1):34. doi: 10.1186/s13071-019-3290-5. PubMed PMID: 30646934; PubMed Central PMCID: PMCPMC6332526.

60. Garms R, Walsh JF, Davies JB. Studies on the reinvasion of the Onchocerciasis Control Programme in the Volta River Basin by Simulium damnosum s.I. with emphasis on the south-western areas. Tropenmed Parasitol. 1979;30(3):345–62. Epub 1979/09/01. PubMed PMID: 575581.

61. Le Berre R, Garms R, Davies JB, Walsh JF, Philippon B, Johnson CG, et al. Displacements of Simulium damnosum and strategy of control against onchocerciasis [and discussion]. Philos Trans R Soc Lond B Biol Sci. 1979;287(1022):277–88. Epub 1979/11/20. doi: 10.1098/rstb.1979.0061. PubMed PMID: 43521.

62. Baker RH, Guillet P, Seketeli A, Poudiougo P, Boakye D, Wilson MD, et al. Progress in controlling the reinvasion of windborne vectors into the western area of the Onchocerciasis Control Programme in West Africa [and discussion]. Philos Trans R Soc Lond B Biol Sci. 1990;328(1251):731–47, discussion 47-50. Epub 1990/06/30. doi: 10.1098/rstb.1990.0141. PubMed PMID: 1976266.

63. Guillet P. Long distance migrations of blackflies and onchocerciasis transmission. In Dadzie Y, Neira M, Hopkins D. Final report of the Conference on the Eradicability of Onchocerciasis. Filaria J. 2003;2(2):53–8.

64. Remme JH. Research for control: the onchocerciasis experience. Trop Med Int Health. 2004;9(2):243–54. PubMed PMID: 15040562.

65. Boatin B. The Onchocerciasis Control Programme in West Africa (OCP). Ann Trop Med Parasitol. 2008;102(Suppl 1):13–7.

66. Wiratsudakul A, Suparit P, Modchang C. Dynamics of Zika virus outbreaks: an overview of mathematical modeling approaches. PeerJ. 2018;6:e4526. Epub 2018/03/30. doi: 10.7717/peerj.4526. PubMed PMID: 29593941; PubMed Central PMCID: PMCPMC5866925.

67. Gambhir M, Michael E. Complex ecological dynamics and eradicability of the vector borne macroparasitic disease, lymphatic filariasis. PLoS One. 2008;3(8):e2874. Epub 2008/08/22. doi: 10.1371/journal.pone.0002874. PubMed PMID: 18716676; PubMed Central PMCID: PMCPMC2518518.

68. Gambhir M, Singh BK, Michael E. The Allee effect and elimination of neglected tropical diseases: a mathematical modelling study. Adv Parasitol. 2015;87:1–31. Epub 2015/03/15. doi: 10.1016/bs.apar.2014.12.001. PubMed PMID: 25765192.

69. Stolk WA, Stone C, de Vlas SJ. Modelling lymphatic filariasis transmission and control: modelling frameworks, lessons learned and future directions. Adv Parasitol. 2015;87:249–91. Epub 2015/03/15. doi: 10.1016/bs.apar.2014.12.005. PubMed PMID: 25765197.

70. Riley S. Large-scale spatial-transmission models of infectious disease. Science. 2007;316(5829):1298–301. Epub 2007/06/02. doi: 10.1126/science.1134695. PubMed PMID: 17540894.

71. Basáñez M-G, Walker M, Turner HC, Coffeng LE, De Vlas SJ, Stolk WA. River blindness: mathematical models for control and elimination. In: Basanez MG, Anderson RM, editors. Mathematical Models for Neglected Tropical Diseases: Essential tools for control and elimination, part B. Advances in Parasitology. 94. New York: Elsevier; 2016.

72. Prost A, Hervouet JP, Thylefors B. Epidemiologic status of onchocerciasis. Bull World Health Organ. 1979;57(4):655–62.

73. Dadzie Y, Amazigo UV, Boatin BA, Seketeli A. Is onchocerciasis elimination in Africa feasible by 2025: a perspective based on lessons learnt from the African control programmes. Infect Dis Poverty. 2018;7(1):63. doi: 10.1186/s40249-018-0446-z. PubMed PMID: 29966535; PubMed Central PMCID: PMCPMC6029117.

74. World Health Organization, African Programme for Onchocerciasis Control. Conceptual and operational framework of onchocerciasis elimination with ivermectin treatment. Geneva: African Programme for Onchocerciasis Control; 2010.

75. Agoua H, Alley ES, Hougard J-M, Akpoboua KLB, Boatin B, Sékétéli A. Études entomologiques de post-traitement dans le programme de lutte contre l’onchocercose en Afrique de l’Ouest / Procedure of definitive cessation of larviciding in the Onchocerciasis Control Programme in West Africa : entomological post-control studies. Parasite. 1995;2:281–8. doi: 10.1051/parasite/1995023281.

76. Bottomley C, Isham V, Vivas-Martinez S, Kuesel AC, Attah SK, Opoku NO, et al. Modelling neglected tropical diseases diagnostics: the sensitivity of skin snips for Onchocerca volvulus in near elimination and surveillance settings. Parasit Vectors. 2016;9(1):343. doi: 10.1186/s13071-016-1605-3. PubMed PMID: 27301567; PubMed Central PMCID: PMCPMC4908809.

77. Organisation mondiale de la Santé, Programme de Lutte contre l’Onchocercose dans la Région du Bassin de la Volta, Prost A, Prod’hon J. Le diagnostic parasitologique de l’onchocercose : revue critique des méthodes en usage. 1977 Contract No.: OCP/STAC6.4.

78. Prost A, Prod’hon J. Le diagnostic parasitologique de l’onchocercose : revue critique des méthodes en usage. Médecine Tropicale. 1978;38(5):519–32.

79. Anderson RM, May RM. Helminth infections of humans: mathematical models, population dynamics, and control. Adv Parasitol. 1985;24:1–101. Epub 1985/01/01. doi: 10.1016/s0065-308x(08)60561-8. PubMed PMID: 3904343.

80. Basáñez Mg, Churcher TS, Grillet ME. Onchocerca-Simulium interactions and the population and evolutionary biology of Onchocerca volvulus. Adv Parasitol. 2009;68:263–313. doi: 10.1016/S0065-308X(08)00611-8. PubMed PMID: 19289198.

81. Duerr HP, Raddatz G, Eichner M. Control of onchocerciasis in Africa: threshold shifts, breakpoints and rules for elimination. Int J Parasitol. 2011;41(5):581–9. Epub 2011/01/25. doi: 10.1016/j.ijpara.2010.12.009. PubMed PMID: 21255577.

82. Turner HC, Walker M, Churcher TS, Osei-Atweneboana MY, Biritwum NK, Hopkins A, et al. Reaching the London Declaration on Neglected Tropical Diseases goals for onchocerciasis: an economic evaluation of increasing the frequency of ivermectin treatment in Africa. Clin Infect Dis. 2014;59(7):923–32. doi: 10.1093/cid/ciu467. PubMed PMID: 24944228; PubMed Central PMCID: PMCPMC4166981.

83. World Health Organization, Onchocerciasis Control Programme in the Volta River Basin Area. Criteria for the definition of “onchocerciasis freed area” and a tolerable level of its transmission. 1977.

84. WHO Expert Committee on Onchocerciasis Control (1993 : Geneva Switzerland), World Health Organization. Onchocerciasis and its control : report of a WHO Expert Committee on Onchocerciasis Control. Geneva: World Health Organization; 1995. 103 p. 85.

85. Rodríguez-Pérez MA, Katholi CR, Hassan HK, Unnasch TR. Large-scale entomologic assessment of Onchocerca volvulus transmission by poolscreen PCR in Mexico. Am J Trop Med Hyg. 2006;74(6):1026–33. PubMed PMID: 16760515.

86. Marino S, Hogue IB, Ray CJ, Kirschner DE. A methodology for performing global uncertainty and sensitivity analysis in systems biology. J Theor Biol. 2008;254(1):178–96.

87. Disney RH, Boreham PF. Blood gorged resting blackflies in Cameroon and evidence of zoophily in Simulium damnosum. Trans R Soc Trop Med Hyg. 1969;63(2):286–7. Epub 1969/01/01. doi: 10.1016/0035-9203(69)90163-1. PubMed PMID: 5794461.

88. Toé L, Merriweather A, Unnasch TR. DNA probe-based classification of Simulium damnosum s. l.-borne and human-derived filarial parasites in the Onchocerciasis Control Program area. Am J Trop Med Hyg. 1994;51(5):676–83. Epub 1994/11/01. PubMed PMID: 7985761.

89. Cheke RA, Denke AM. Anthropophily, zoophily and roles in onchocerciasis transmission of the Djodji form of Simulium sanctipauli and S. squamosum in a forest zone of Togo. Trop Med Parasitol. 1988;39(2):123–7. PubMed PMID: 3175468.

90. Disney RH. Observations on chicken-biting blackflies in Cameroon with a discussion of parous rates of Simulium damnosum. Ann Trop Med Parasitol. 1972;66(1):149–58. Epub 1972/03/01. doi: 10.1080/00034983.1972.11686807. PubMed PMID: 5021565.

91. Thompson BH. Studies on the attraction of Simulium damnosum s.l. (Diptera: Simuliidae) to its hosts. III. Experiments with animal-baited traps. Tropenmed Parasitol. 1977;28(2):226–8. Epub 1977/06/01. PubMed PMID: 888186.

92. Philippon B. Étude de la transmission d’Onchocerca volvulus (Leuckart, 1893) (Nematoda, Onchocercidae) par Simulium damnosum Theobald, 1903 (Diptera: Simuliidae) en Afrique tropicale. Trav Doc ORSTOM. 1977;63:308.

93. Lamberton PH, Cheke RA, Walker M, Winskill P, Crainey JL, Boakye DA, et al. Onchocerciasis transmission in Ghana: the human blood index of sibling species of the Simulium damnosum complex. Parasit Vectors. 2016;9(1):432. Epub 2016/08/09. doi: 10.1186/s13071-016-1703-2. PubMed PMID: 27494934; PubMed Central PMCID: PMCPMC4975878.

94. McCulloch K, Golding N, McVernon J, Goodwin S, Tomko M. Ensemble model for estimating continental-scale patterns of human movement: a case study of Australia. Sci Rep. 2021;11(1):4806. Epub 2021/02/28. doi: 10.1038/s41598-021-84198-6. PubMed PMID: 33637816; PubMed Central PMCID: PMCPMC7910534.

95. Lakwo T, Garms R, Wamani J, Tukahebwa EM, Byamukama E, Onapa AW, et al. Interruption of the transmission of Onchocerca volvulus in the Kashoya-Kitomi focus, western Uganda by long-term ivermectin treatment and elimination of the vector Simulium neavei by larviciding. Acta Trop. 2017;167:128–36. doi: 10.1016/j.actatropica.2016.12.029. PubMed PMID: 28034767.

96. World Health Organization, Onchocerciasis Control Programme in West Africa. Twenty years of onchocerciasis control in West Africa: review of the work of the Onchocerciasis Control Programme in West Africa from 1974 to 1994. Geneva, Switzerland: World Health Organization, 1997.

97. Jacob BG, Loum D, Lakwo TL, Katholi CR, Habomugisha P, Byamukama E, et al. Community-directed vector control to supplement mass drug distribution for onchocerciasis elimination in the Madi mid-North focus of Northern Uganda. PLoS Negl Trop Dis. 2018;12(8):e0006702. Epub 2018/08/28. doi: 10.1371/journal.pntd.0006702. PubMed PMID: 30148838; PubMed Central PMCID: PMCPMC6128654.

98. Jacob B, Loum D, Munu D, Lakwo T, Byamukama E, Habomugisha P, et al. Optimization of slash and clear community-directed control of Simulium damnosum sensu stricto in northern Uganda. Am J Trop Med Hyg. 2021. Epub 2021/01/13. doi: 10.4269/ajtmh.20-1104. PubMed PMID: 33432900; PubMed Central PMCID: PMCPMC8045649.

99. Raimon S, Lakwo TL, Sebit WJ, Siewe Fodjo JN, Alinda P, Carter JY, et al. “Slash and clear”, a community-based vector control method to reduce onchocerciasis transmission by Simulium sirbanum in Maridi, South Sudan: a prospective study. Pathogens. 2021;10(10):1329.

100. Stolk WA, Blok DJ, Hamley JID, Cantey PT, de Vlas SJ, Walker M, et al. Scaling-down mass ivermectin treatment for onchocerciasis elimination: modeling the impact of the geographical unit for decision making. Clin Infect Dis. 2021;72(Suppl 3):S165–S71. Epub 2021/04/29. doi: 10.1093/cid/ciab238. PubMed PMID: 33909070; PubMed Central PMCID: PMCPMC8201558.

101. Koala L, Grant WN, McCulloch K, Hedtke SM, Kuesel AC, Boakye D, editors. CORNTD Research Links: Delineation of transmission zones to improve the evidence base for stop MDA decisions and reduce the risk of resurgence 2021.

