## Supplementary material for "Impact of human movement between hypo- and hyperendemic areas on sustainability of elimination of *Onchocerca volvulus* transmission": S1 File

### 1 Model equations and parameters

The intensity of *O. volvulus* infection changes over time and with host age, as described by the following system of partial differential equations (PDEs). The model describes, with respect to time ($t$) and host age ($a$), the number of adult worms in human host of sex $s$ resident in patch $i$ ($W_{s,i}$), the number of microfilariae in human host of sex $s$ resident in patch $i$ ($M_{s,i}$) and the number of infective larvae in the black fly population in patch $i$ biting human hosts of sex $s$ ($L_{s,i}$). Note that the terms for $W_{s,i}$ , $M_{s,i}$ and $L_{s,i}$ depend on time and age but these dependencies are omitted from our notation for simplicity. Table S1 provides a detailed description of the variables and parameters in the patch model.

##### Table S1: Partial differential equations for the 2- patch model

| Equations for patch 1 | PDE |
| --- | --- |
| $\frac{\boldsymbol{\partial}\boldsymbol{W}_{\boldsymbol{s}\mathbf{,}\boldsymbol{1}}}{\boldsymbol{\partial}\boldsymbol{t}}\mathbf{+}\frac{\boldsymbol{\partial}\boldsymbol{W}_{\boldsymbol{s}\mathbf{,1}}}{\boldsymbol{\partial}\boldsymbol{a}}\mathbf{=}\left\{ \boldsymbol{1}\mathbf{-}\boldsymbol{f}_{\boldsymbol{1}}\left( \boldsymbol{a} \right) \right\}\boldsymbol{m}_{\boldsymbol{1}}\boldsymbol{\beta}_{\boldsymbol{1}}\boldsymbol{\Omega}_{\boldsymbol{s}}\left( \boldsymbol{a}\mathbf{-}\boldsymbol{p} \right)\boldsymbol{\Pi}_{\boldsymbol{H}}\left[ \boldsymbol{L}_{\boldsymbol{s}\mathbf{,}\boldsymbol{1}} \right]\mathbb{L}_{\boldsymbol{1}}\left( \boldsymbol{t} \right)$ $\mathbf{+}\boldsymbol{f}_{\boldsymbol{1}}\left( \boldsymbol{a} \right)\boldsymbol{d}_{\boldsymbol{12}}\boldsymbol{m}_{\boldsymbol{2}}\boldsymbol{\beta}_{\boldsymbol{2}}\boldsymbol{\Omega}_{\boldsymbol{s}}\left( \boldsymbol{a}\mathbf{-}\boldsymbol{p} \right)\boldsymbol{\Pi}_{\boldsymbol{H}}\left[ \boldsymbol{L}_{\boldsymbol{s}\mathbf{,}\boldsymbol{2}} \right]\mathbb{L}_{\boldsymbol{2}}\left( \boldsymbol{t} \right)\mathbf{-}\boldsymbol{\sigma}_{\boldsymbol{W}\boldsymbol{1}}\boldsymbol{W}_{\boldsymbol{s}\mathbf{,}\boldsymbol{1}}$ | (S.1) |
| $\frac{\boldsymbol{\partial}\boldsymbol{M}_{\boldsymbol{s}\mathbf{,}\boldsymbol{1}}}{\boldsymbol{\partial}\boldsymbol{t}}\mathbf{+}\frac{\boldsymbol{\partial}\boldsymbol{M}_{\boldsymbol{s}\mathbf{,}\boldsymbol{1}}}{\boldsymbol{\partial}\boldsymbol{a}}\mathbf{=}\boldsymbol{\Delta}\boldsymbol{W}_{\boldsymbol{s}\mathbf{,}\boldsymbol{1}}\mathbf{-}\boldsymbol{\sigma}_{\boldsymbol{M}\boldsymbol{1}}\boldsymbol{M}_{\boldsymbol{s}\mathbf{,}\boldsymbol{1}}$ | (S.2) |
| $\frac{\boldsymbol{\partial}\boldsymbol{L}_{\boldsymbol{s}\mathbf{,}\boldsymbol{1}}}{\boldsymbol{\partial}\boldsymbol{t}}\mathbf{+}\frac{\boldsymbol{\partial}\boldsymbol{L}_{\boldsymbol{s}\mathbf{,}\boldsymbol{1}}}{\boldsymbol{\partial}\boldsymbol{a}}\mathbf{=}\left\{ \boldsymbol{1}\mathbf{-}\boldsymbol{f}_{\boldsymbol{1}}\left( \boldsymbol{a} \right) \right\}\boldsymbol{\beta}_{\boldsymbol{1}}\boldsymbol{\Omega}_{\boldsymbol{s}}\left( \boldsymbol{a} \right)\boldsymbol{\Pi}_{\boldsymbol{V}}\left[ \boldsymbol{M}_{\boldsymbol{s}\mathbf{,}\boldsymbol{1}} \right]\boldsymbol{M}_{\boldsymbol{s}\mathbf{,}\boldsymbol{1}}\mathbf{+}\boldsymbol{\beta}_{\boldsymbol{1}}\boldsymbol{\Omega}_{\boldsymbol{s}}\left( \boldsymbol{a} \right)\boldsymbol{f}_{\boldsymbol{2}}\left( \boldsymbol{a} \right)\boldsymbol{d}_{\boldsymbol{21}}\boldsymbol{\Pi}_{\boldsymbol{V}}\left[ \boldsymbol{M}_{\boldsymbol{s}\mathbf{,}\boldsymbol{2}} \right]\boldsymbol{M}_{\boldsymbol{s}\mathbf{,}\boldsymbol{2}}$  $\mathbf{-}\boldsymbol{v}_{\boldsymbol{12}}\boldsymbol{L}_{\boldsymbol{s}\mathbf{,}\boldsymbol{1}}\mathbf{+}\boldsymbol{v}_{\boldsymbol{21}}\boldsymbol{L}_{\boldsymbol{s}\mathbf{,}\boldsymbol{2}}\mathbf{-}\boldsymbol{\sigma}_{\boldsymbol{L}\boldsymbol{1}}\boldsymbol{L}_{\boldsymbol{s}\mathbf{,}\boldsymbol{1}}$ | (S.3) |
| Equations for patch 2 |  |
| $\frac{\boldsymbol{\partial}\boldsymbol{W}_{\boldsymbol{s}\mathbf{,}\boldsymbol{2}}}{\boldsymbol{\partial}\boldsymbol{t}}\mathbf{+}\frac{\boldsymbol{\partial}\boldsymbol{W}_{\boldsymbol{s}\mathbf{,}\boldsymbol{2}}}{\boldsymbol{\partial}\boldsymbol{a}}\mathbf{=}\left\{ \boldsymbol{1}\mathbf{-}\boldsymbol{f}_{\boldsymbol{2}}\left( \boldsymbol{a} \right) \right\}\boldsymbol{m}_{\boldsymbol{2}}\boldsymbol{\beta}_{\boldsymbol{2}}\boldsymbol{\Omega}_{\boldsymbol{s}}\left( \boldsymbol{a}\mathbf{-}\boldsymbol{p} \right)\boldsymbol{\Pi}_{\boldsymbol{H}}\left[ \boldsymbol{L}_{\boldsymbol{s}\mathbf{,}\boldsymbol{2}} \right]\mathbb{L}_{\boldsymbol{2}}\left( \boldsymbol{t} \right)$  $\mathbf{+}\boldsymbol{f}_{\boldsymbol{2}}\left( \boldsymbol{a} \right)\boldsymbol{d}_{\boldsymbol{21}}\boldsymbol{m}_{\boldsymbol{1}}\boldsymbol{\beta}_{\boldsymbol{1}}\boldsymbol{\Omega}_{\boldsymbol{s}}\left( \boldsymbol{a}\mathbf{-}\boldsymbol{p} \right)\boldsymbol{\Pi}_{\boldsymbol{H}}\left[ \boldsymbol{L}_{\boldsymbol{s}\mathbf{,}\boldsymbol{1}} \right]\mathbb{L}_{\boldsymbol{1}}\left( \boldsymbol{t} \right)\mathbf{-}\boldsymbol{\sigma}_{\boldsymbol{W}\boldsymbol{2}}\boldsymbol{W}_{\boldsymbol{s}\mathbf{,}\boldsymbol{2}}$ | (S.4) |
| $\frac{\boldsymbol{\partial}\boldsymbol{M}_{\boldsymbol{s}\mathbf{,}\boldsymbol{2}}}{\boldsymbol{\partial}\boldsymbol{t}}\mathbf{+}\frac{\boldsymbol{\partial}\boldsymbol{M}_{\boldsymbol{s}\mathbf{,}\boldsymbol{2}}}{\boldsymbol{\partial}\boldsymbol{a}}\mathbf{=}\boldsymbol{\Delta}\boldsymbol{W}_{\boldsymbol{s}\mathbf{,}\boldsymbol{2}}\mathbf{-}\boldsymbol{\sigma}_{\boldsymbol{M}\boldsymbol{2}}\boldsymbol{M}_{\boldsymbol{s}\mathbf{,}\boldsymbol{2}}$ | (S.5) |
| $\frac{\boldsymbol{\partial}\boldsymbol{L}_{\boldsymbol{s}\mathbf{,}\boldsymbol{2}}}{\boldsymbol{\partial}\boldsymbol{t}}\mathbf{+}\frac{\boldsymbol{\partial}\boldsymbol{L}_{\boldsymbol{s}\mathbf{,}\boldsymbol{2}}}{\boldsymbol{\partial}\boldsymbol{a}}\mathbf{=}\left\{ \boldsymbol{1}\mathbf{-}\boldsymbol{f}_{\boldsymbol{2}}\left( \boldsymbol{a} \right) \right\}\boldsymbol{\beta}_{\boldsymbol{2}}\boldsymbol{\Omega}_{\boldsymbol{s}}\left( \boldsymbol{a} \right)\boldsymbol{\Pi}_{\boldsymbol{V}}\left[ \boldsymbol{M}_{\boldsymbol{s}\mathbf{,}\boldsymbol{2}} \right]\boldsymbol{M}_{\boldsymbol{s}\mathbf{,}\boldsymbol{2}}\mathbf{+}\boldsymbol{\beta}_{\boldsymbol{2}}\boldsymbol{\Omega}_{\boldsymbol{s}}\left( \boldsymbol{a} \right)\boldsymbol{f}_{\boldsymbol{1}}\left( \boldsymbol{a} \right)\boldsymbol{d}_{\boldsymbol{12}}\boldsymbol{\Pi}_{\boldsymbol{V}}\left[ \boldsymbol{M}_{\boldsymbol{s}\mathbf{,}\boldsymbol{1}} \right]\boldsymbol{M}_{\boldsymbol{s}\mathbf{,}\boldsymbol{1}}$  $\mathbf{-}\boldsymbol{v}_{\boldsymbol{21}}\boldsymbol{L}_{\boldsymbol{s}\mathbf{,}\boldsymbol{2}}\mathbf{+}\boldsymbol{v}_{\boldsymbol{12}}\boldsymbol{L}_{\boldsymbol{s}\mathbf{,}\boldsymbol{1}}\mathbf{-}\boldsymbol{\sigma}_{\boldsymbol{L}\boldsymbol{2}}\boldsymbol{L}_{\boldsymbol{s}\mathbf{,}\boldsymbol{2}}$ | (S.6) |
| where  $\boldsymbol{\sigma}_{\boldsymbol{Li}}\mathbf{=}\frac{\boldsymbol{a}_{\boldsymbol{H}}}{\boldsymbol{g}}\mathbf{+}\boldsymbol{\sigma}_{\boldsymbol{L}_{\boldsymbol{0}}}\mathbf{+}\boldsymbol{\mu}_{\boldsymbol{V}}\mathbf{+}\boldsymbol{\alpha}_{\boldsymbol{V}}\boldsymbol{M}_{\boldsymbol{s}\mathbf{,}\boldsymbol{i}}$ | (S.7) |
| and  $\mathbb{L}_{\boldsymbol{i}}\left( \boldsymbol{t} \right)\mathbf{=}\sum_{\boldsymbol{s}} \boldsymbol{\int}\boldsymbol{\rho}\left( \boldsymbol{a} \right)\boldsymbol{\Omega}_{\boldsymbol{s}}\left( \boldsymbol{a} \right)\boldsymbol{L}_{\boldsymbol{s}\mathbf{,}\boldsymbol{i}}\left( \boldsymbol{a} \right)\boldsymbol{da}\mathbf{.}$ | (S.8) |

We solve the system of PDEs by discretizing the human population into one year age groups and incorporating an annual rate of aging, converting the model into a system of ordinary differential equations (ODEs). The system of ODEs were solved in MATLAB[1] using the *ode23s* solver with 100 integration timesteps per year and reporting solutions at yearly intervals over an 80 year period.

Patch model code can be accessed through <https://github.com/shedtke/Onchocerciasis_patch_model>. Supplementary File S1 Text contains further details including the patch model equations for an arbitrary number of patches and an explanation of how prevalence was derived.

##### Table S2: Definitions of variables and parameters in the patch model

| **Symbol** | **Definition** | **Value (*distribution*)** | **Reference** |
| --- | --- | --- | --- |
| **Parasite in human host** | | | |
| $W_{s,i}$ | Average number of adult worms in human host of sex $s$ and age $a$ in patch $i$ | - |  |
| $M_{s,i}$ | Average number of microfilariae in human host of sex $s$ and age $a$ in patch $i$ | - |  |
| $\delta_{H}\left( L\left( t \right) \right)$ | Proportion of infective larvae developing into adult worms within the human host | $\frac{\delta_{H0}+\delta_{H\infty}c_{H}m\beta L\left( t \right)}{1+c_{H}m\beta L\left( t \right)}$ | [2] |
| $\delta_{H_{0}}$ | Proportion of L3 larvae developing to adult worms within the human host when $m\beta L\left( t \right)\to0$ | 0.0854; *Uniform(0.0712 - 0.16)* | [3, 4] |
| $\delta_{H_{\infty}}$ | Proportion of L3 larvae developing to adult worms within the human host when $m\beta L\left( t \right)\to\infty$ | 0.00299; *Uniform(0.002 – 0.004)* | [3, 4] |
| $c_{H}$ | Severity of transmission intensity-dependent parasite establishment within the human host | $5.86\times{10}^{-3}$ year per L3 larva | [4] |
| $p$ | Prepatent period (from infection with L3 larvae to presence of detectable microfilariae in the skin) | $2$ years | [3] |
| $\mu_{H}$ | Per capita death rate of human hosts | $0.04$ year${}^{-1}$ | [3] |
| $\sigma_{Wi}$ | Per capita mortality rate of adult worms | $0.1$ year${}^{-1}$ ; *Uniform(0.0909 - 0.1111)* | [4, 5] |
| $\sigma_{Mi}$ | Per capita mortality rate of microfilariae | $0.8$ year${}^{-1}$; *Uniform(0.5 - 1)* | [4, 6] |
| $\Phi$ | Mating probability of adult worms | Assumed to equal 1 | [3] |
| $F$ | Fecundity rate of adult female worms | 0.67 | [3] |
| $\Delta$ | Birth rate of microfilariae, assumes half adult worm population is female | $\phi F/2$. | [3] |
| $a_{m}$ | Maximum recorded human age in the reference population of northern Cameroon | 80 years | [3] |
| $\rho_{s}$ | Proportion of host population in sex group $s$ | $\rho_{F}=0.45$; $\rho_{M}=0.55$ | [3] |
| $f_{i}\left( a \right)$ | Proportion of the human host population in age group $a$ resident in patch $i$ that travel | Values considered with range $\left[ 0,0.5 \right]$ |  |
| $\{1-f_{i}\left( a \right)\}$ | Proportion of the human host population in age group $a$ resident in patch $i$ that *do not* travel | Depends on $f_{i}\left( a \right)$ values |  |
| $d_{ij}$ | Fraction of time the population in patch $i$ that travel spends in patch $j$ | For a 2-patch model:  $d_{12}$ = $d_{21}$ = 1. |  |
| $f_{ij}\left( a \right)$ | Fraction of time human hosts in age group *a* resident in patch *i* spend in patch *j* | $f_{i}\left( a \right)\times d_{ij}$ |  |
| **Parasite in simuliid vector** | | | |
| $L_{s,i}$ | Average number of infective larvae in the black fly population biting human hosts of sex $s$ and age $a$ in patch $i$ | - |  |
| $\mathbb{L}_{i}$ | Average number of infective larvae in the black fly population in patch $i$ | - |  |
| $h$ | Human blood index: proportion of blood meals taken on humans | 0.3; *Uniform(0.3,0.99)* | [2, 3, 7] [4][8] |
| $g$ | Average duration between consecutive blood meals | 0.0096 year. *Uniform(0.0086, 0.011)* | [2-4] |
| $\beta_{i}$ | Biting rate of black flies in patch $i$ | $\beta_{i}=\frac{h}{g}$ | [2] |
| $ABR_{i}$ | Annual biting rate in patch $i$ | $ABR_{1}=1,000;$ *Uniform(100, 1100)*  $ABR_{2}=4,000;$*Uniform(2000, 16000)* |  |
| $m_{i}$ | Ratio of vectors to humans within patch $i$ | $m_{i}=\frac{ABR_{i}}{\beta_{i}}$ |  |
| $\delta_{V}\left[ M_{s}\left( t,a \right) \right]$ ($\Pi_{V}$) | Proportion of microfilariae developing into to the infective stage within the vector, per bite | $\frac{\delta_{v0}}{1+c_{V}M_{s}\left( t,a \right)}$ | [2] |
| $\sigma_{Li}$ | Mortality rate of infective larvae; incorporates infection induced and background mortality | $\frac{a_{H}}{g}+\sigma_{L0}+\mu_{V}+\alpha_{V}M_{s}\left( t,a \right)$ | [3] |
| $\delta_{V0}$ | Proportion of microfilariae developing to L3 within vectors when $M_{s}\left( t,a \right)\to0$, per bite | 0.0207 [2] (0.005 if ignoring density dependence in $\Pi_{V}$ [3] with $c_{V}=0$) | [2, 3] |
| $c_{V}$ | Severity of density-dependent limitation of larval development within (savannah) vectors | 0.0148 (0 if ignoring density dependence in $\Pi_{V}$ with  $\delta_{V0}=0.005$) | [2] |
| $a_{H}$ | Proportion of infective, L3 larvae shed per bite | 0.5; *Uniform(0.5 - 0.8)* | [3, 9, 10] |
| $\sigma_{L0}$ | Per capita death rate of L3 larvae within the vector | 104 year${}^{-1}$ *Uniform(52,104)* | [3, 4, 11] |
| $\mu_{V}$ | Per capita death rate of uninfected blackflies | 52 year${}^{-1}$ *Uniform(26,52)* | [2, 11] |
| $\alpha_{V}$ | Parasite-induced death rate of infected blackflies | 0.597 year${}^{-1}$ per microfilaria | [3, 4] |
| $\Omega_{s}\left( a \right)$ | Age- and sex- dependent exposure to blackflies | $\left\{ \begin{aligned} E_{s}\gamma_{s}E_{0}, a<a^{'} \\ E_{s}\gamma_{s}e^{-\alpha_{s}(a-a^{'})}, a>a' \end{aligned} \right.$ | [3] |
| $E_{s}$ | Sex-specific exposure to blackfly bites | $E_{F}=0.9$; $E_{M}=1.08$ | [3] |
| $E_{0}$ | Fraction of exposure at age 0 in relation to that at age $a'$ from which exposure changes with age | 0.10; in Cameroon $a'$ was estimated to be = 0 | [3] |
| $\gamma_{s}$ | Normalization factors to ensure that the distribution of bites among age groups sums to 1 | $\gamma_{F}=0.548$; $\gamma_{M}=1.154$ | [3] |
| $\alpha_{s}$ | Age-specific change in contact rate with vectors for human hosts of sex $s$ | $\alpha_{F}=-0.023$; $\alpha_{M}=0.007$ | [3] |
| $v_{ij}$ | Migration rate per year of black flies from patch $i$ to patch $j$ | Set to 0 for this paper |  |
| $ATP_{i}$ | Annual transmission potential in patch $i$ | $ATP_{i}=m_{i}\beta_{i}\mathbb{L}_{i}\left( t \right)$ |  |
| **Parasite prevalence** | | | |
| $k_{M}\left( M_{i}\left( t \right) \right)$ | Inverse measure of the degree of overdispersion in the distribution of skin microfilariae among hosts, as a function of the mean microfilarial load | $\frac{k_{0}M_{i}\left( t \right)}{1+k_{1}M_{i}\left( t \right)}$ | [12] |
| $k_{0}$ | Parameters of the relationship between $k_{M}$ and skin microfilarial load | 0.013 | [12] |
| $k_{1}$ | Parameters of the relationship between $k_{M}$ and skin microfilarial load | 0.025 | [12] |
| $\pi_{i}$ | Microfilarial prevalence in patch $i$ | $1-\left[ 1+\frac{M_{i}\left( t \right)}{k_{M}\left( M_{i}\left( t \right) \right)} \right]^{-k_{M}\left( M_{i}\left( t \right) \right)}$ | [12] |

##### Table S3: Partial differential equations for a patch model with a number of patches ≥2

The below system of equations describes the mean parasite burden at each life stage of *Onchocerca volvulus* in the $i^{th}$ patch. Note these equations describe the general case where $\left\{ i,j \right\}\in\left\{ 1,\ldots, n \right\}$ for $j\neq i$ and where *n* is the total number of patches in the system.

| General equations for a patch model | PDE |
| --- | --- |
| $\frac{\boldsymbol{\partial}\boldsymbol{W}_{\boldsymbol{s}\mathbf{,i}}}{\boldsymbol{\partial}\boldsymbol{t}}\mathbf{+}\frac{\boldsymbol{\partial}\boldsymbol{W}_{\boldsymbol{s}\mathbf{,i}}}{\boldsymbol{\partial}\boldsymbol{a}}\mathbf{=}\left\{ \boldsymbol{1}\mathbf{-}\boldsymbol{f}_{\boldsymbol{i}}\left( \boldsymbol{a} \right) \right\}\boldsymbol{m}_{\boldsymbol{i}}\boldsymbol{\beta}_{\boldsymbol{i}}\boldsymbol{\Omega}_{\boldsymbol{s}}\left( \boldsymbol{a}\mathbf{-}\boldsymbol{p} \right)\boldsymbol{\Pi}_{\boldsymbol{H}}\left[ \boldsymbol{L}_{\boldsymbol{s}\mathbf{,}\boldsymbol{i}} \right]\mathbb{L}_{\boldsymbol{i}}\left( \boldsymbol{t} \right)$ $\mathbf{+}\sum_{\boldsymbol{j}}^{\boldsymbol{n}} \boldsymbol{f}_{\boldsymbol{i}}\left( \boldsymbol{a} \right)\boldsymbol{d}_{\boldsymbol{ij}}\boldsymbol{m}_{\boldsymbol{j}}\boldsymbol{\beta}_{\boldsymbol{j}}\boldsymbol{\Omega}_{\boldsymbol{s}}\left( \boldsymbol{a}\mathbf{-}\boldsymbol{p} \right)\boldsymbol{\Pi}_{\boldsymbol{H}}\left[ \boldsymbol{L}_{\boldsymbol{s}\mathbf{,j}} \right]\mathbb{L}_{\boldsymbol{j}}\left( \boldsymbol{t} \right)\mathbf{-}\boldsymbol{\sigma}_{\boldsymbol{Wi}}\boldsymbol{W}_{\boldsymbol{s}\mathbf{,i}}$ | (S.9) |
| $\frac{\boldsymbol{\partial}\boldsymbol{M}_{\boldsymbol{s}\mathbf{,i}}}{\boldsymbol{\partial}\boldsymbol{t}}\mathbf{+}\frac{\boldsymbol{\partial}\boldsymbol{M}_{\boldsymbol{s}\mathbf{,i}}}{\boldsymbol{\partial}\boldsymbol{a}}\mathbf{=}\boldsymbol{\Delta}\boldsymbol{W}_{\boldsymbol{s}\mathbf{,i}}\mathbf{-}\boldsymbol{\sigma}_{\boldsymbol{Mi}}\boldsymbol{M}_{\boldsymbol{s}\mathbf{,i}}$ | (S.10) |
| $\frac{\boldsymbol{\partial}\boldsymbol{L}_{\boldsymbol{s}\mathbf{,i}}}{\boldsymbol{\partial}\boldsymbol{t}}\mathbf{+}\frac{\boldsymbol{\partial}\boldsymbol{L}_{\boldsymbol{s}\mathbf{,i}}}{\boldsymbol{\partial}\boldsymbol{a}}\mathbf{=}\left\{ \boldsymbol{1}\mathbf{-}\boldsymbol{f}_{\boldsymbol{i}}\left( \boldsymbol{a} \right) \right\}\boldsymbol{\beta}_{\boldsymbol{i}}\boldsymbol{\Omega}_{\boldsymbol{s}}\left( \boldsymbol{a} \right)\boldsymbol{\Pi}_{\boldsymbol{V}}\left[ \boldsymbol{M}_{\boldsymbol{s}\mathbf{,}\boldsymbol{i}} \right]\boldsymbol{M}_{\boldsymbol{s}\mathbf{,}\boldsymbol{i}}\mathbf{+}\boldsymbol{\beta}_{\boldsymbol{i}}\boldsymbol{\Omega}_{\boldsymbol{s}}\left( \boldsymbol{a} \right)\sum_{\boldsymbol{j}}^{\boldsymbol{n}} \boldsymbol{f}_{\boldsymbol{j}}\left( \boldsymbol{a} \right)\boldsymbol{d}_{\boldsymbol{ji}}\boldsymbol{\Pi}_{\boldsymbol{V}}\left[ \boldsymbol{M}_{\boldsymbol{s}\mathbf{,}\boldsymbol{j}} \right]\boldsymbol{M}_{\boldsymbol{s}\mathbf{,}\boldsymbol{j}}$  $\mathbf{-}\sum_{\boldsymbol{j}}^{\boldsymbol{n}} \boldsymbol{v}_{\boldsymbol{ij}}\boldsymbol{L}_{\boldsymbol{s}\mathbf{,}\boldsymbol{i}}\mathbf{+}\sum_{\boldsymbol{j}}^{\boldsymbol{n}} \boldsymbol{v}_{\boldsymbol{ji}}\boldsymbol{L}_{\boldsymbol{s}\mathbf{,}\boldsymbol{j}}\mathbf{-}\boldsymbol{\sigma}_{\boldsymbol{Li}}\boldsymbol{L}_{\boldsymbol{s}\mathbf{,}\boldsymbol{i}}$ | (S.11) |
| where  $\boldsymbol{\sigma}_{\boldsymbol{Li}}\mathbf{=}\frac{\boldsymbol{a}_{\boldsymbol{H}}}{\boldsymbol{g}}\mathbf{+}\boldsymbol{\sigma}_{\boldsymbol{L}_{\boldsymbol{0}}}\mathbf{+}\boldsymbol{\mu}_{\boldsymbol{V}}\mathbf{+}\boldsymbol{\alpha}_{\boldsymbol{V}}\boldsymbol{M}_{\boldsymbol{s}\mathbf{,}\boldsymbol{i}}$ | (S.12) |
| and  $\mathbb{L}_{\boldsymbol{i}}\left( \boldsymbol{t} \right)\mathbf{=}\sum_{\boldsymbol{s}} \boldsymbol{\int}\boldsymbol{\rho}\left( \boldsymbol{a} \right)\boldsymbol{\Omega}_{\boldsymbol{s}}\left( \boldsymbol{a} \right)\boldsymbol{L}_{\boldsymbol{s}\mathbf{,}\boldsymbol{i}}\left( \boldsymbol{a} \right)\boldsymbol{da}\mathbf{.}$ | (S.13) |

##### References

1.	The MathWorks Inc., *MATLAB version 9.7 (R2019b)*. Natick, Massachusetts.

2.	Basáñez, M.-G. and M. Boussinesq, *Population biology of human onchocerciasis.* Philos Trans R Soc Lond B Biol Sci, 1999. **354**(1384): p. 809-826.

3.	Filipe, J.A., et al., *Human infection patterns and heterogeneous exposure in river blindness.* Proc Natl Acad Sci U S A, 2005. **102**(42): p. 15265-70.

4.	Basáñez, M.-G., et al., *Transmission intensity and the patterns of Onchocerca volvulus infection in human communities.* Am J Trop Med Hygiene, 2002. **67**(6): p. 669-679.

5.	Plaisier, A.P., et al., *The reproductive lifespan of Onchocerca volvulus in West African savanna.* Acta Tropica, 1991. **48**(4): p. 271-284.

6.	Duke, B.O., *The population dynamics of Onchocerca volvulus in the human host.* Trop Med Parasitol, 1993. **44**(2): p. 61-68.

7.	Basáñez, M.G., et al., *Density-dependent processes in the transmission of human onchocerciasis: relationship between the numbers of microfilariae ingested and successful larval development in the simuliid vector.* Parasitology, 1995. **110**(4): p. 409-427.

8.	Lamberton, P.H.L., et al., *Onchocerciasis transmission in Ghana: the human blood index of sibling species of the Simulium damnosum complex.* Parasit Vectors, 2016. **9**(1).

9.	Renz, A., *Studies on the dynamics of transmission of onchocerciasis in a Sudan-savanna area of North Cameroon III. Infection rates of the Simulium vectors and Onchocerca volvulus transmission potentials.* Ann Trop Med Parasitol, 1987. **81**(3): p. 239-52.

10.	Duke, B.O., *Studies on factors influencing the transmission of onchocerciasis. 8. The escape of infective Onchocerca volvulus larvae from feeding 'forest' Simulium damnosum.* Ann Trop Med Parasitol, 1973. **67**(1): p. 95-9.

11.	Basáñez, M.G., et al., *River blindness: mathematical models for control and elimination.* Adv Parasitol, 2016. **94**: p. 247-341.

12.	Turner, H.C., et al., *Modelling the impact of ivermectin on River Blindness and its burden of morbidity and mortality in African Savannah: EpiOncho projections.* Parasit Vectors, 2014. **7**(1): p. 241.

13.	Stolk, W.A., et al., *Required duration of mass ivermectin treatment for onchocerciasis elimination in Africa: a comparative modelling analysis.* Parasit Vectors, 2015. **8**(1).

14.	Turner, H.C., et al., *Uncertainty surrounding projections of the long-term impact of ivermectin treatment on human onchocerciasis.* PLoS Negl Trop Dis, 2013. **7**(4): e2169.

### 2 Supplemental Figures

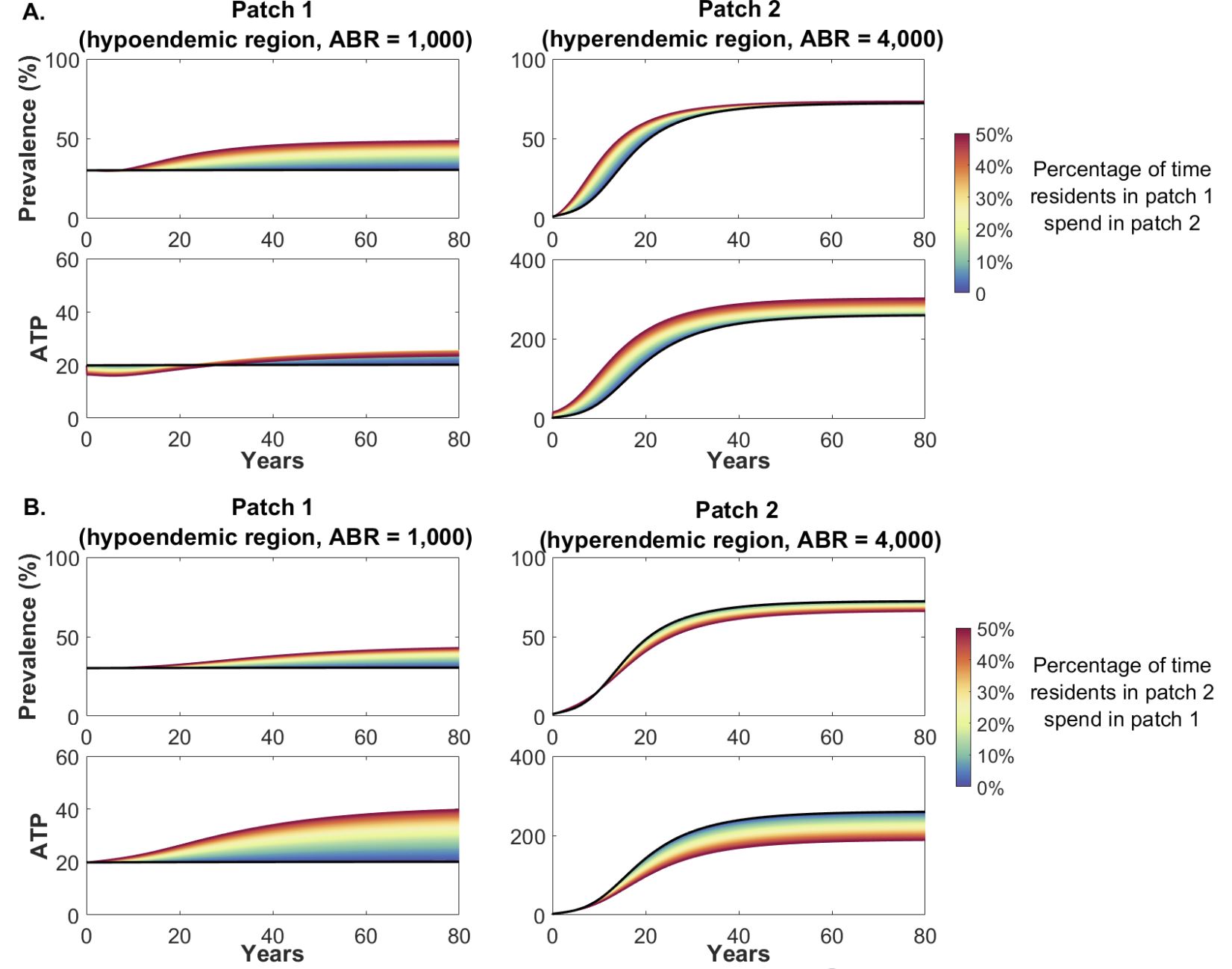

##### Figure S1. Change in microfilariae prevalence among population (aged ≥5) and annual transmission potential (ATP) in patch 1 and patch 2 due to human hosts aged 20 to 50 years (35% of the population) travelling between the patches post-MDAi.

Note the difference in scale for the ATP of patch 1 and patch 2. **A.** Effect of individuals living in the hypoendemic region (patch 1) spending a percentage of their time in the post-MDAi hyperendemic region (patch 2). In this scenario the parameter $f_{12}$ is varied while assuming that residents of patch 2 do not travel to patch 1 ($f_{21}=0$). **B.** Effect of individuals living in patch 2 spending a percentage of their time in the hypoendemic region (patch 1). In this scenario the parameter $f_{21}$ is varied while assuming that residents of patch 1 do not travel to patch 2 ($f_{12}=0$). Solid black lines indicate prevalence and ATP if the two patches are not coupled; i.e., when residents of one area do not spend time in the other.

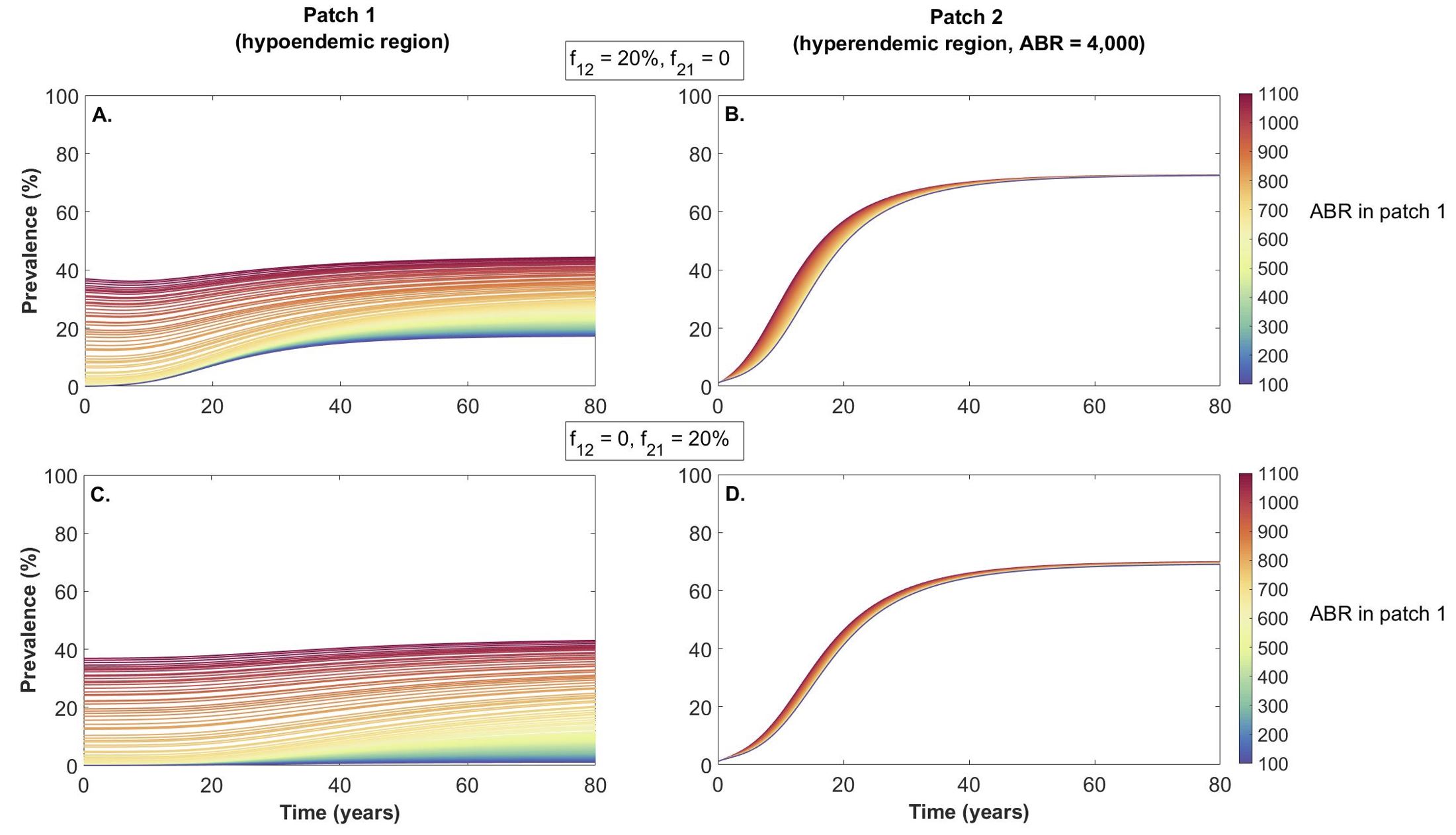

##### Figure S2. Impact of variation in the Annual Biting Rate (ABR) of blackfly vectors in a hypoendemic region (patch 1) on microfilariae prevalence (in ages 5 and above) when coupled to a hyperendemic region (patch 2) post-MDAi.

Patch 1 is initialized by running the model assuming the patch is isolated to equilibrium for each ABR_1_ value; Patch 2 is initialized with <1.4% mf prevalence.

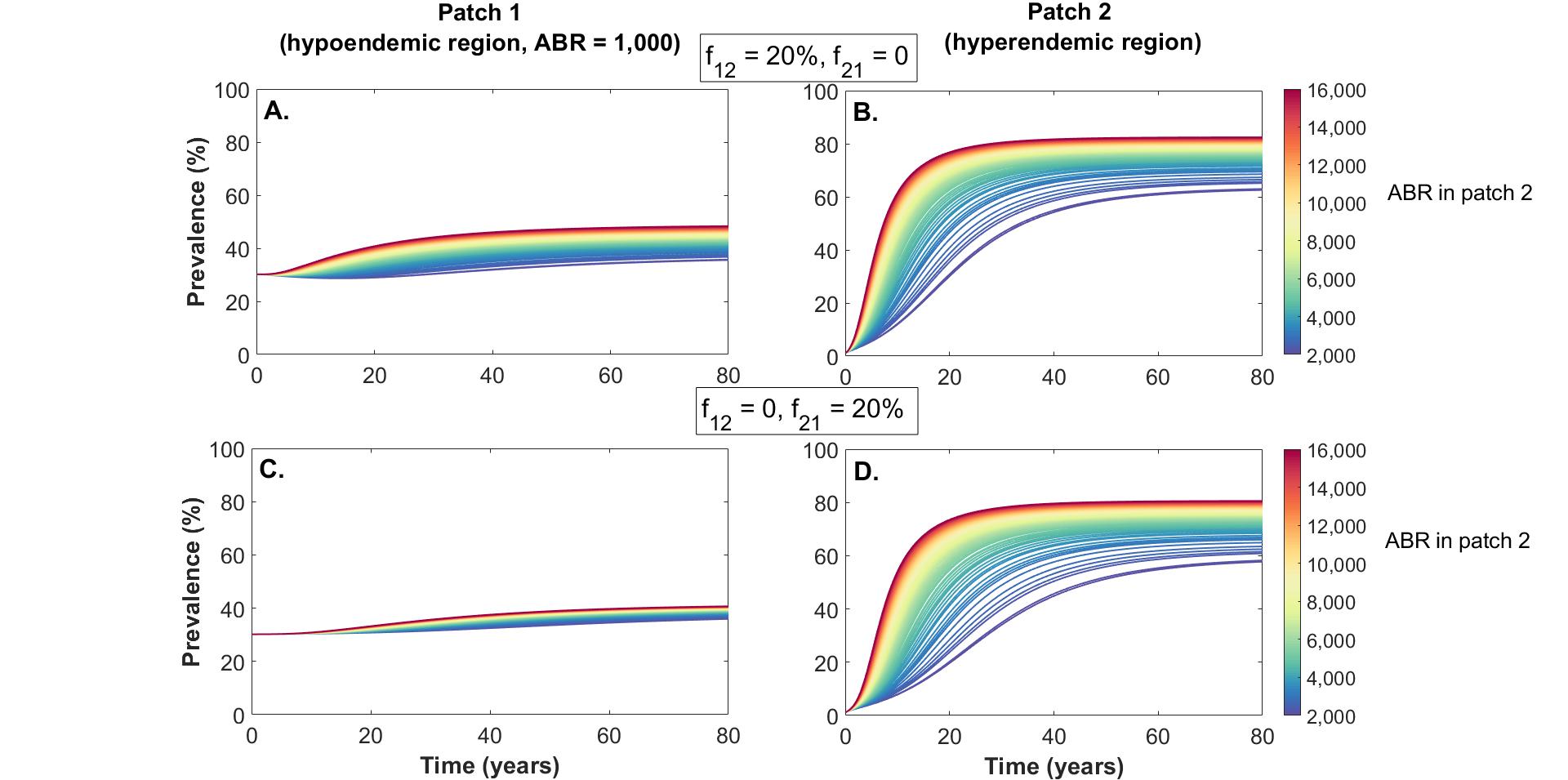

##### Figure S3. Impact of variation in the ABR in a hyperendemic region (patch 2) has on mf prevalence (in ages 5 and above) when coupled to a hypoendemic region (patch 1) post-MDAi.

Patch 1 is initialized by running the model assuming the patch is isolated to equilibrium (with ABR_1_= 1,000); Patch 2 is initialized with <1.4% mf prevalence.

### 3 Can host movement sustain transmission in a region which has an insufficient ABR to sustain ongoing transmission if isolated?

Threshold biting rates (TBRs) and transmission breakpoints have been studied for existing models of *Onchocerca volvulus* transmission and utilized to understand conditions required to sustain transmission in isolated regions. Here, we explored (via simulations) the impact of coupling a region with an ABR too low to sustain transmission on its own with a region with hyperendemic prevalence. For the set of parameters provided in Table S2, if the ABR within an isolated patch is less than $\approx750$ then the prevalence of infection will decline over time while an ABR within an isolated patch greater than $\approx750$ can sustain transmission within that hypoendemic area.

Therefore, we set the ABR in patch 1 to be 600 and the ABR in patch 2 to be 4,000. Both patches were initialized with a mf prevalence $<1.4\%$, and simulated for a burn in period of 200 years assuming the patches are isolated, and then simulated for a further 80 years with different levels of coupling (extent of unidirectional or bidirectional human movement). Fig s5 shows the mf prevalence at the end of these 80 years.

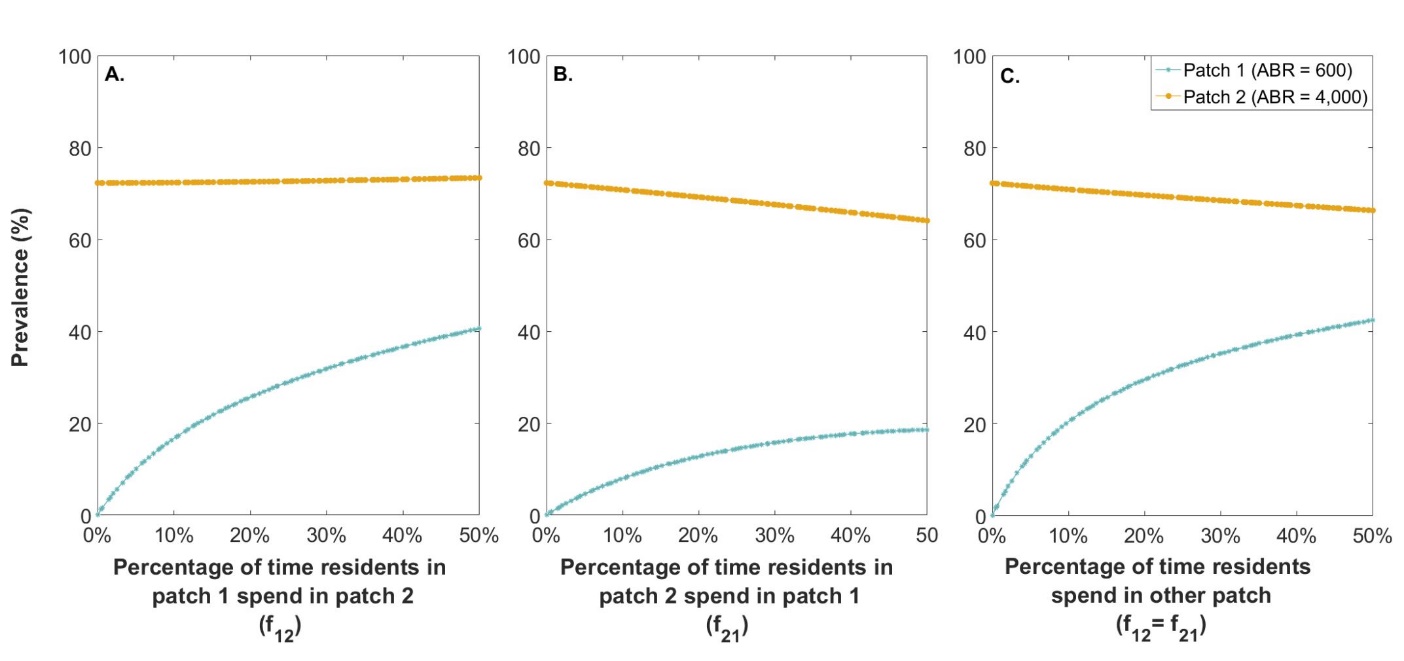

##### Figure S54 Long-term prevalence of mf in the human hosts aged 5 years and older in the presence of human movement.

Prevalence for each patch was estimated from the model after 80 years. Note when the movement parameter equals zero on the horizontal axis ($f_{12}$, $f_{21}$ and both $f_{12}$& $f_{21}$ for graphs A, B and C respectively), the two patches are isolated.

Figure S4 illustrates that when a hypoendemic region with an insufficient ABR to sustain transmission on its own is coupled (via human movement) to a hyperendemic region, transmission can be sustained and indeed prevalence will increase. The resulting long-term prevalence in the hypoendemic region will depend on the strength and direction of human movement occurring between the two regions.

### 4 Impact of the human blood index on the relationship between ABR and modeled prevalence

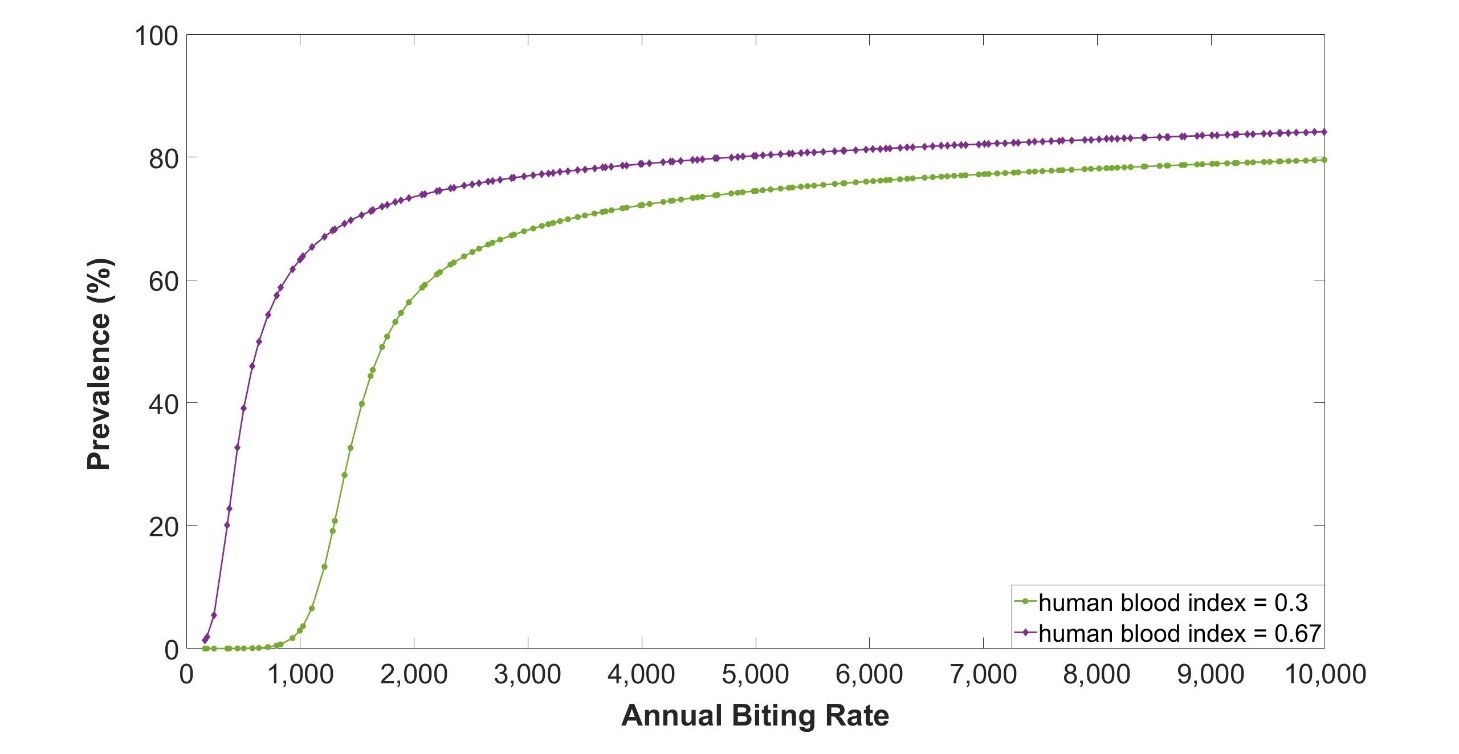

##### Figure S5: Impact of human blood index (h) on the relationship between the annual biting rate and modeled prevalence in ages 5 years and over.

The threshold biting rate will be lower if the human blood index is increased while holding all other parameters stable, or in other words ongoing transmission would be possible with lower ABR.
